# Antisense oligonucleotide depletion of *CCDC146* is a broad-spectrum therapeutic strategy for ALS

**DOI:** 10.1101/2024.03.30.24305115

**Authors:** Sai Zhang, Tobias Moll, Jasper Rubin-Sigler, Sharon Tu, Shuya Li, Enming Yuan, Menghui Liu, Marriane King, Elham Alhathli, Connie Treanor, Alison Twelvetrees, Marcel Weinreich, Sam Bonsall, Calum Harvey, Sarah Gornall, Allan Shaw, Johanna Ganssauge, Akshay Bhinge, Cleide Dos Santos Souza, Laura Ferraiuolo, Eran Hornstein, Ophir Shalem, Anna Avila, Tatyana Shelkovnikova, Charlotte H. van Dijk, Ilia S. Timpanaro, Kevin P. Kenna, Jianyang Zeng, Philip van Damme, Philip S. Tsao, Pamela J. Shaw, Justin K. Ichida, Johnathan Cooper-Knock, Michael P. Snyder

## Abstract

Amyotrophic lateral sclerosis (ALS) is a heritable and incurable disease defined by the degeneration of motor neurons (MNs), yet the genetics of ALS remain partially understood. Using a genomic deep learning-powered whole-genome analysis of 6,715 ALS patients, we identify four rare noncoding variants associated with patient survival, including chr7:76,009,472:C>T which is linked to a 70.6% reduction in survival. Genetic editing of this variant into iPSC-derived MNs increases *CCDC146* expression and exacerbates ALS-specific phenotypes including TDP-43 mislocalization. We reveal that CCDC146 was located within the basal body of primary cilia in human MNs, and that cilia structure and function is impaired by *CCDC146* overexpression but is restored by its depletion. Suppressing *CCDC146* using an antisense oligonucleotide (ASO) completely rescues ALS-specific survival defects in neurons derived from both sporadic and familial patients, and it extends survival and reverses TDP-43 pathology in an aggressive ALS mouse model. Taken together, CCDC146 is a new modifier of ALS survival that acts via the primary cilia of MNs. ASO targeting of *CCDC146* is a potential therapeutic approach for both sporadic and genetic forms of ALS, particularly because congenital absence of CCDC146 is well tolerated.

## Introduction

Amyotrophic lateral sclerosis (ALS) is a devastating neurodegenerative disease^1^ characterized by the progressive loss of motor neurons (MNs), ultimately leading to respiratory failure and death. ALS survival is markedly heterogeneous: the median survival is between 20 and 48 months, but 10-20% of ALS patients live longer than 10 years^2^. This heterogeneity persists even between patients who share the same monogenic risk factor^3^. A wide range of genetic variations and genes have been associated with ALS risk^4^, but a lack of understanding of genetic factors influencing ALS progression is hindering the development of therapies to prolong patient survival.

Conventional genetic analyses, such as genome-wide association studies (GWAS) and rare variant association studies (RVAS) are likely to be underpowered because of limited functional characterization of genetic variants^5, 6^. This limitation is pronounced for noncoding variants, which comprise the majority of human genetic material and complex disease heritability^7^. Understanding the biological function of noncoding variants requires specific cellular context because of their involvement in the transcriptional regulation that defines cell identity^8^. Moreover, *rare* variants pose additional challenges in traditional studies of variant function, such as the expression quantitative trait locus (eQTL) analysis^9, 10^, which rely on prevalent cohort-level evidence. Recent advances in sequence-to-function (seq2func) deep learning models^11–16^, also known as genomic deep learning models, provide a powerful alternative method to predict at high-resolution the functional effects of noncoding variants and bypassing the requirement for individual-level data.

In this study, we developed a gDL-powered genetic analysis framework (**Fig. 1**) and performed thus-far the largest (*n* = 6,715) rare variant analysis for ALS survival, with a focus on the noncoding genetic variants with a functional effect in human MNs. We discovered four novel rare noncoding variants that are associated with an average reduction in ALS survival of more than 50%. We introduced the top scored variant chr7:76,009,472:C>T (where score indicates the probability of an effect on gene expression within MN) into MNs derived from an ALS patient. We confirmed an effect of chr7:76,009,472:C>T on *CCDC146* expression, and an exacerbation of phenotypes associated with ALS including neuronal survival, dendritic regression, TDP-43 localization, and disruption of primary cilia structure and function. Depletion of *CCDC146 in vitro* using an antisense oligonucleotide (ASO) rescued ALS-specific survival deficits in patient-derived neurons, irrespective of the genetic background or of baseline *CCDC146* expression, and without significant toxicity. ASO targeting of murine *Ccdc146* extended survival by over 50% in an aggressive ALS mouse model caused by TDP-43 mislocalization^17^. Indeed the ASO reversed TDP-43 mislocalization and prevented MN loss in the mouse. We localized CCDC146 within the basal body of primary cilia^18^ of MNs and discovered that the structure and function of primary cilia is inversely proportional to CCDC146 expression. Cilia dysfunction has been associated with ALS^19^, but our work positions *CCDC146* and MN cilia as a therapeutic target with relevance across the spectrum of ALS patients.

**Fig. 1.**
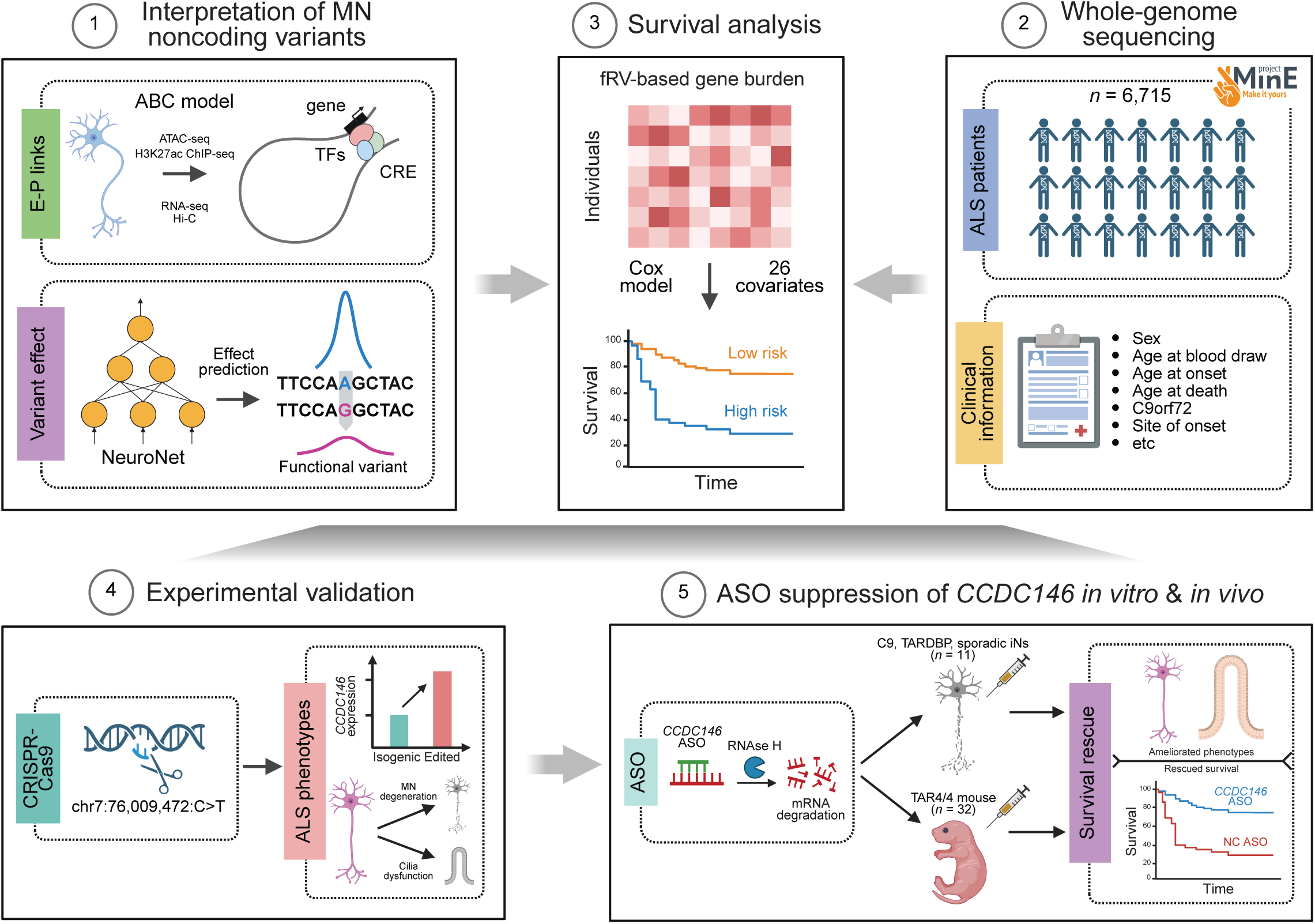
Schematic of the study design. Leveraging epigenomic and transcriptomic profiling of iPSC-derived human motor neurons (iMNs), we first identified enhancer-promoter (E-P) interactions using the Activity-by-Contact (ABC)^20^ model (**Panel 1**, top). Next, we developed a genomic deep learning model^12, 14^ named NeuroNet to prioritize functional noncoding variants that could significantly alter MN epigenomes (**Panel 1**, bottom). Using the Project MinE^29^ whole genome sequencing (WGS) data (**Panel 2**), we identified risk variants and genes modulating ALS survival (**Panel 3**) via the Cox proportional-hazards (PH) model, in which we collapsed all functional rare noncoding variants (predicted by NeuroNet) to their target genes (predicted by the ABC model). To validate our discovery, we introduced the top candidate variant chr7:76,009,472:C>T into patient-derived iMNs using CRISPR-Cas9 (**Panel 4**, left), and verified its regulatory effects on *CCDC146* expression and ALS-associated phenotypes (**Panel 4**, right), including cell survival, neurite growth, and TDP-43 mislocalization. Finally, we designed ASOs suppressing *CCDC146* (**Panel 5**, left). We demonstrated that *CCDC146* ASOs rescued ALS-associated survival deficits in patient-derived neurons across different genetic backgrounds, and improved motor phenotypes, survival, and MN loss in an aggressive ALS mouse model (**Panel 5**, right). iPSC, induced pluripotent stem cell; CRE, cis-regulatory element; TF, transcription factor; fRV, functional rare variant; ASO, antisense oligonucleotide; RNase H, Ribonuclease H.

## Results

### Overview of the study design

We developed a functional genomics-informed genetic analysis pipeline (**Fig. 1**) to improve discovery power and interpretability. Briefly, we first predicted the functional impacts of noncoding variants on gene regulation in human MNs utilizing a gDL. Functional variants were then mapped to their target genes based on the enhancer-promoter (E-P) interactions constructed within MNs. Informed by these variant-to-gene maps, a gene burden-based survival analysis was performed using 6,025 high-quality ALS genomes (out of 6,715 genomes), and pinpointing survival-associated rare variants and corresponding genes. Next, we measured the effect of the top variant chr7:76,009,472:C>T, and changes in expression of its target gene *CCDC146,* in iPSC-derived MNs to determine the underlying biology. Finally, we designed ASOs targeting *CCDC146* to test whether CCDC146 depletion can rescue disease-associated survival defects across different ALS genetic backgrounds, both *in vitro* and i*n vivo*. This framework is generic and can be broadly applied to study rare variant genetics of other complex diseases.

### Constructing the regulome and predicting noncoding variant effects in human motor neurons

To characterize the transcriptional regulation in human motor neurons, we mapped MN E-P interactions by applying the activity-by-contact (ABC) model^20^ (**Supplementary Fig. 1A**) to the transcriptomic (RNA-seq) and epigenomic (ATAC-seq, H3K27ac ChIP-seq, and Hi-C) profiling data of iPSC-derived MNs (iMN) derived from neurologically normal donors^21^ (**Supplementary Fig. 1B** and **1C**, **Supplementary Table 1**, **Supplementary Results**; **Methods**). A large proportion of MN E-P links (89.0%) were cell-type-specific (**Supplementary Fig. 1D**), and were relatively enriched with long-range interactions (**Supplementary Fig. 1E**, **Supplementary Results**). As expected, transcription factors (TFs) and genes involved in MN E-P links were enriched with biological pathways known to be critical for MN function (**Supplementary Fig. 1F-1H** and **2A**, **Supplementary Results**).

Next, we sought to examine the function of noncoding variants specifically within MNs. The activity of epigenomic features, such as chromatin accessibility and histone modifications, varies widely across cell types and contexts, yet much of this regulation is encoded in the underlying DNA sequence^11, 12, 14^. To learn this epigenome grammar in MNs, we developed a genomic deep learning model^12, 22^, named NeuroNet, to predict chromatin accessibility and key histone marks (H3K27ac, H3K4me1, and H3K4me3) in iMNs directly from the reference DNA sequence (**Fig. 2A**; **Methods**). After training (**Methods**), NeuroNet accurately predicted these epigenomic features, achieving a mean area under receiver operating characteristic curve (AUROC) of 0.851 (SD = 0.10; **Fig. 2B**) and a mean area under precision-recall curve (AUPRC) of 0.853 (SD = 0.063; **Fig. 2C**).

**Fig. 2.**
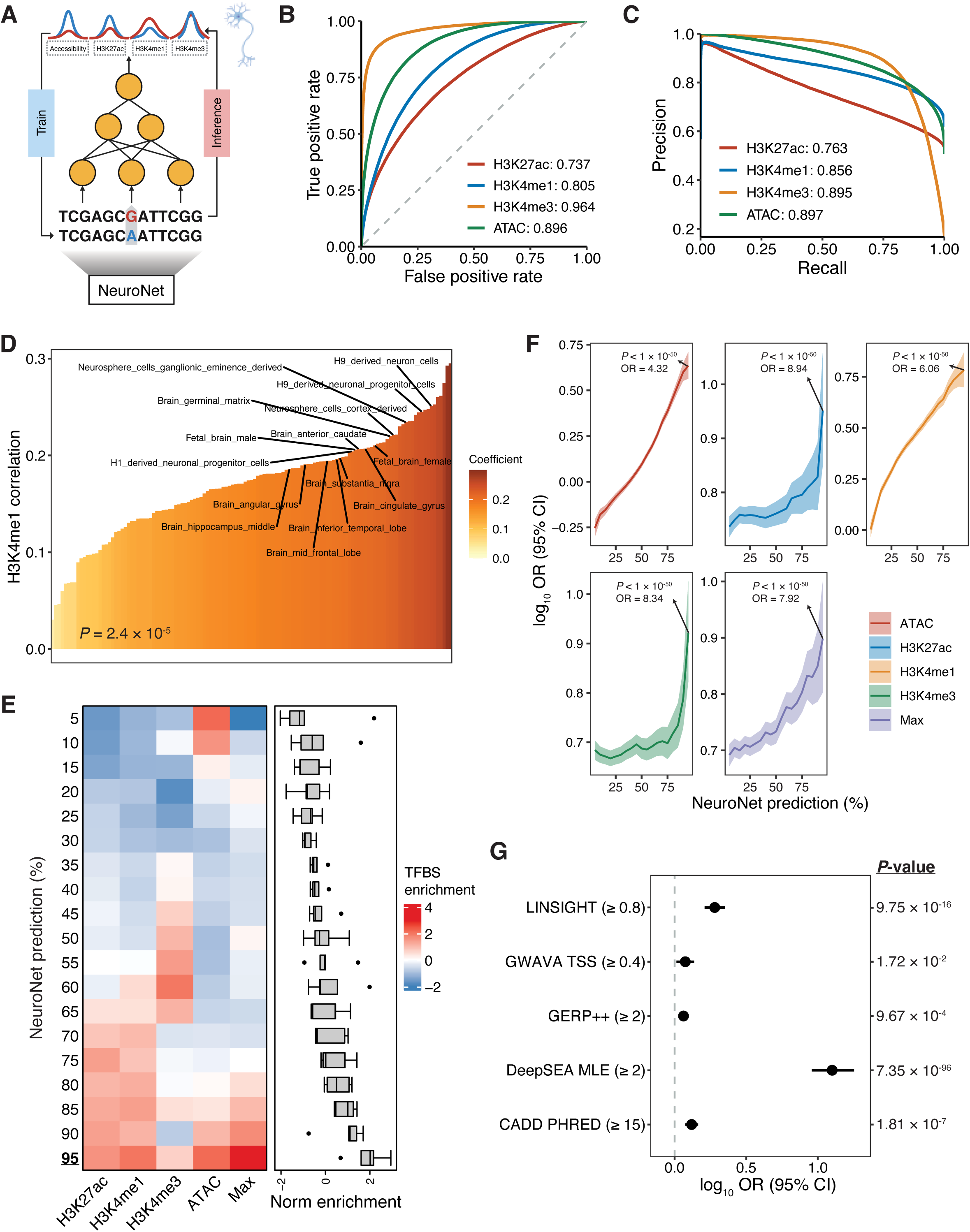
NeuroNet predicts the noncoding variant effects in MNs. (**A**) Schematics of the NeuroNet model. (**B-C**) Predictive performance of NeuroNet evaluated based on the receiver operating characteristic (ROC) curve (**B**) and precision-recall curve (**C**). The area under curve (AUC) scores were calculated for different epigenomic features and metrics. The dashed gray line indicates the random prediction. (**D**) Correlation between NeuroNet and DeepSEA predictions for H3K4me1. *P*-value by two-sided Fisher’s exact test. Neuron- and brain-related tissues and cell types are highlighted. PCC, Pearson correlation coefficient. (**E**) Enrichment of TFBS-disrupting variants within NeuroNet-prioritized functional variants (various cutoffs applied). Enrichment was estimated by *t*-statistics. The box plot center line, limits, and whiskers represent the median, quartiles, and 1.5x interquartile range (IQR), respectively. TFBS, transcription factor binding site. (**F**) Enrichment of NeuroNet-prioritized functional variants (various cutoffs applied) within MN CREs. OR is annotated in the solid line and 95% CI is represented in the colored area. Statistics of the 95% cutoff were highlighted. *P*-value by two-sided Fisher’s exact test. OR, odds ratio; CI, confidence interval. (**G**) Enrichment of functional or deleterious variants predicted by traditional approaches within NeuroNet-prioritized functional variants (cutoff = 0.06). *P*-value by two-sided Fisher’s exact test. Dot and error bar indicate OR and 95% CI, respectively. MLE, Mean -log e-value.

We then used NeuroNet to characterize the functional effects of noncoding variants in MNs by querying the difference in predicted epigenomic features between reference and alternative alleles (**Fig. 2A**; **Methods**). Differing from traditional approaches in modeling variant effects^23^, this *in-silico* strategy requires no individual-level data, which greatly increases its flexibility and generalizability. NeuroNet outputs a score that defines the probability of a specific variant altering a given epigenomic feature. To benchmark our variant effect prediction, we compared NeuroNet predictions with those of DeepSEA^12^, another deep learning model which has been used to predict 2,002 epigenomic features in various tissues and cell types. As expected, NeuroNet and DeepSEA predictions aligned well in neuron- and brain-related tissues and cell types compared to other cell-types (H3K4me1: *P* = 2.4 x 10^-5^; H3K4me3: *P* = 4.5 x 10^-3^, two-sided Fisher’s exact test; **Fig. 2D** and **Supplementary Fig. 2B**). The alignment was marginally significant for H3K27ac (*P* = 0.059, two-sided Fisher’s exact test; **Supplementary Fig. 2C**), but not significant for chromatin accessibility (*P* > 0.05, two-sided Fisher’s exact test; **Supplementary Fig. 2D**), probability due to the limited number (*n* = 3) of neurons and brains profiled by the DNase I hypersensitive sites sequencing (DNase-seq). In ongoing analyses, to integrate information from all epigenomic features, we used the maximum of all four variant-specific scores to derive the final NeuroNet variant score.

NeuroNet is designed to capture functional consequences of noncoding variants including, but not limited to, TF binding disruption. To aid the biological interpretation of NeuroNet predictions, we examined the overlap between NeuroNet-prioritized variants and variants previously associated with changes in TF binding^24^. We then used this association to determine which NeuroNet score should be used as a ‘cutoff’ to assign ‘functional’ noncoding variants. Specifically, we split NeuroNet prediction scores into percentiles and examined which cutoff produced the strongest enrichment with variants that disrupt TF binding^24^ (**Methods**). Variants with higher NeuroNet scores were more enriched with TF-binding-disrupting variants (**Fig. 2E**). This demonstrates that NeuroNet has captured known TF biology, despite the fact that TF motifs were *not* included in model training. The strongest enrichment with disruption to known TF binding motifs was achieved at the 95% cutoff (NeuroNet score ≈ 0.06; *P* = 5.5 x 10^-4^, *t*-statistics = 3.46, two-sided *t*-test; **Fig. 2E**). Accordingly, we classified variants with a NeuroNet score > 0.06 as ‘functional’ within MN in the remainder of our analyses.

To further examine the functional implication of NeuroNet prediction, we assessed the distribution of prioritized variants across the genome. Notably, we found that MN cis-regularoty elements (CREs), including promoters and enhancers, were enriched with variants prioritized by NeuroNet (odds ratio [OR] > 1.0, two-sided Fisher’s exact test; **Fig. 2F**; **Methods**). As in our TF binding site analysis, this enrichment was positively correlated with the NeuroNet score in all features, and peak enrichment was obtained at the 95% cutoff threshold (**Fig. 2F**).

Finally, we benchmarked our variant effect prediction against multiple widely-used methods, including GERP++^25^, GWAVA^26^, CADD^27^, LINSIGHT^28^, and DeepSEA^12^. By following the suggested cutoffs to define functional or deleterious variants for different approaches, we observed that NeuroNet-defined functional variants were enriched with functional or deleterious variants prioritized by other methods (*P* < 0.05, OR > 1.0, two-sided Fisher’s exact test; **Fig. 2G**; **Methods**). The strongest enrichment was obtained for DeepSEA, probably because the model design is most similar to NeuroNet.

### NeuroNet-powered rare variant analysis for ALS survival

We sought to integrate NeuroNet prediction with ALS whole-genome sequencing (WGS) data to identify rare noncoding variants that impact ALS risk or survival via disrupting transcriptional regulation in MNs.

We performed variant-level and sample-level quality controls (QCs) for 9,600 whole genomes generated in the Project MinE^29^ (Data Freeze 2), resulting in an analysis-ready cohort encompassing 6,025 ALS patients and 2,264 controls (**Methods**). Next, using NeuroNet scores, we identified functional noncoding rare variants (allele frequency [AF] < 0.5% within gnomAD v2.1 non-Finnish European [NFE] samples, allele count [AC] ≥ 3 in the analysis cohort, and NeuroNet score > 0.06) located within MN CREs. We mapped these variants to their target genes based on the ABC-predicted E-P links. A gene-level rare variant burden profile (i.e., the number of variants mapped to each gene) was then constructed for each individual in the QC’ed cohort, ready for downstream genetic association analysis.

We first studied the genetics of ALS risk. We conducted a rare variant burden risk analysis (ALS patients versus healthy controls) using SKAT-O^30, 31^ to quantify the burden of NeuroNet-prioritised variants (**Methods**), with covariates including age, sex, age^2^, sex x age, sequencing platform, and the first 20 principal components to adjust for known confounders. No gene was significantly associated with ALS risk after multiple testing correction, suggesting an underpowered analysis (**Supplementary Table 2**). However, a number of genes that have been linked to ALS risk achieved nominal significance, such as *KIF5A*^32, 33^ (*P* = 0.016) and *NEAT1*^34^ (*P* = 0.033).

Next, we performed a rare variant burden survival analysis using the Cox proportional-hazards (PH) model to measure the effect of NeuroNet-prioritised variants (**Methods**). We included 26 covariates: age, sex, age^2^, sex x age, sequencing platform, *C9orf72* status, and the first 20 principal components, to adjust for known confounders. Noncoding rare variant burden was significantly associated with ALS survival for seven genes after Bonferroni multiple testing correction (adjusted *P* < 0.05; **Fig. 3A** and **3B**, **Supplementary Table 3**), for which no inflation was observed (*λ* = 0.99). The mean survival time for the carriers of these rare variants ranges from 0.99 to 2.46 years, which is 2.79 to 1.32 years shorter than that of the overall cohort (**Fig. 3C**).

**Fig. 3.**
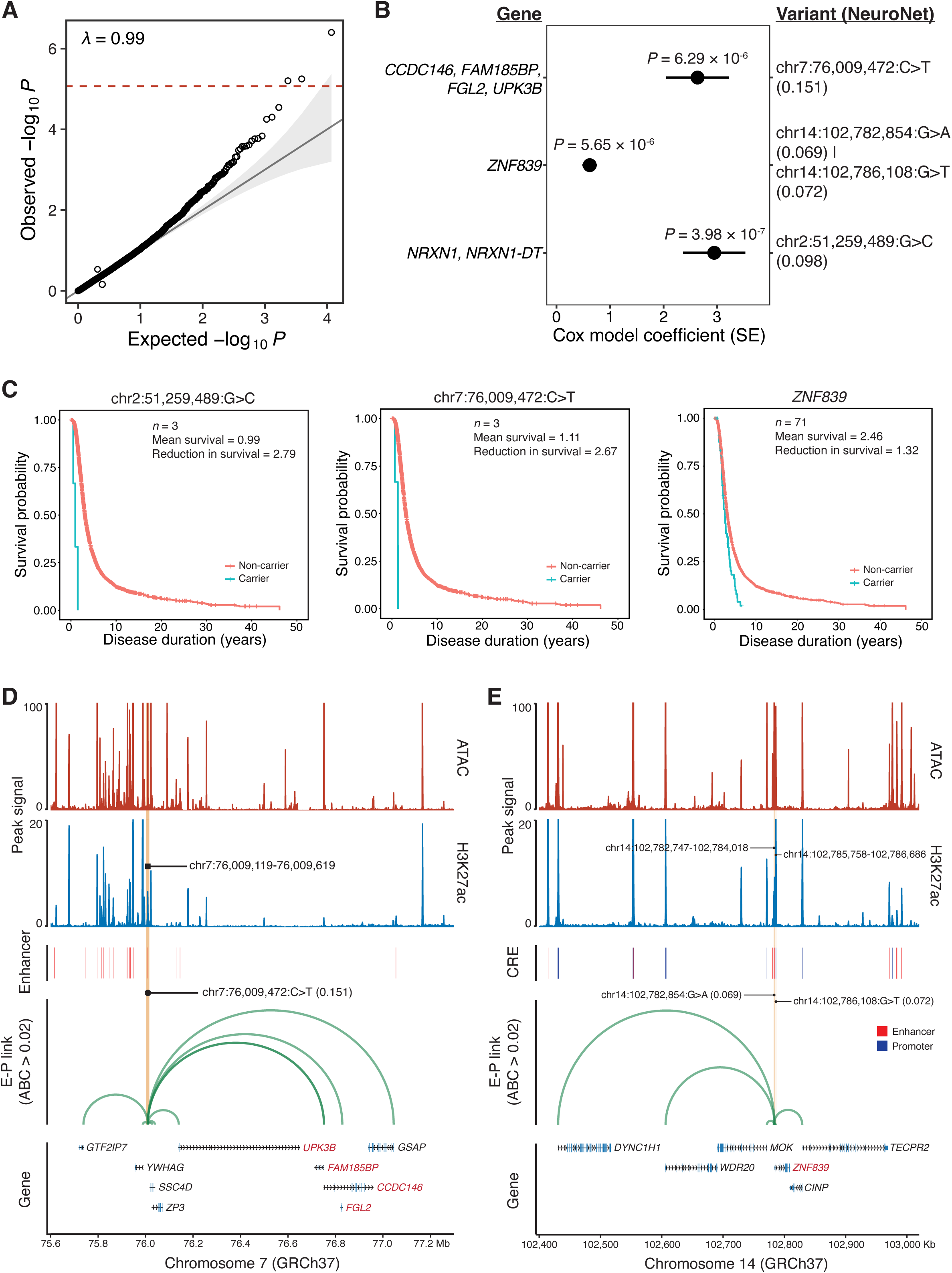
Survival analysis based on rare functional noncoding variants in motor neurons. (**A**) Quantile-quantile (QQ) plot of burden-based survival analysis. Genes sharing the same set of variants were collapsed into groups. The dashed red line indicates the significance cutoff based on the Bonferroni correction. (**B**) Statistics of significant tests. The dot and error bar represent the Cox model coefficient and SE, respectively. SE, standard error. (**C**) Kaplan-Meier (KM) estimator plots for significant tests. Carriers for *ZNF839* variants were defined as individuals having at least one variant from chr14:102,782,854:G>A and chr14:102,786,108:G>T. (**D-E**) Regulatory relationships between noncoding variants and target genes for chr7:76,009,472:C>T (**D**) and *ZNF839* (**E**), respectively. Peak signals from MNs derived from CS14-iPSCs are visualized. Significant genes after Bonferroni correction (adjusted *P* < 0.05) are highlighted in red. CRE, cis-regulatory element; E-P, enhancer-promoter.

Four variants were linked to three gene sets: chr7:76,009,472:C>T resides within an enhancer linked to *CCDC146*, *FAM185BP*, *FGL2*, and *UPK3B* (**Fig. 3D**); chr14:102,782,854:G>A and chr14:102,786,108:G>T are located within an enhancer of *ZNF839* (**Fig. 3E**); chr2:51,259,489:G>C is within the promoter of *NRXN1* (**Supplementary Fig. 3A**). As a benchmark, we performed a standard rare variant burden survival analysis without variant prioritization using NeuroNet (**Methods**), but no gene passed multiple testing correction (**Supplementary Fig. 3B** and **Supplementary Table 4**). These results demonstrate the superiority of our deep learning-based framework in enhancing discovery power and biological interpretation.

Lastly, we were interested in the overlap of rare variant associations between ALS survival and risk. We examined this by testing the enrichment of survival-associated genes within risk-associated genes under different *P*-value cutoffs. Notably, we observed significant gene overlaps across a spectrum of *P*-value cutoffs (*P* < 0.05, two-sided Fisher’s exact test; **Supplementary Fig. 3C**), suggesting an overlapping genetic architecture underlying these two ALS phenotypes within noncoding genomic regions.

### Reproducing chr7:76,009,472:C>T within iMNs alters CCDC146 expression and exacerbates ALS-associated phenotypes

The noncoding variant chr7:76,009,472:C>T achieved the highest NeuroNet score amongst noncoding variants associated with ALS survival (**Fig. 3B**). This variant was linked to four candidate target genes based on predicted E-P links (**Fig. 3D**). To experimentally verify our prediction and determine the regulatory target of chr7:76,009,472:C>T, we utilized CRISPR-Cas9 to reproduce this variant in iPSCs derived from an ALS patient caused by a G4C2-repeat expansion within *C9orf72* (**Fig 4A**; **Methods**), where the unedited cells served as an isogenic control. Of note, we observed extremely rapid disease progression in a patient who carried both a *C9orf72* expansion and chr7:76,009,472:C>T in the Project MinE cohort (**Supplementary Table 5**).

**Fig. 4.**
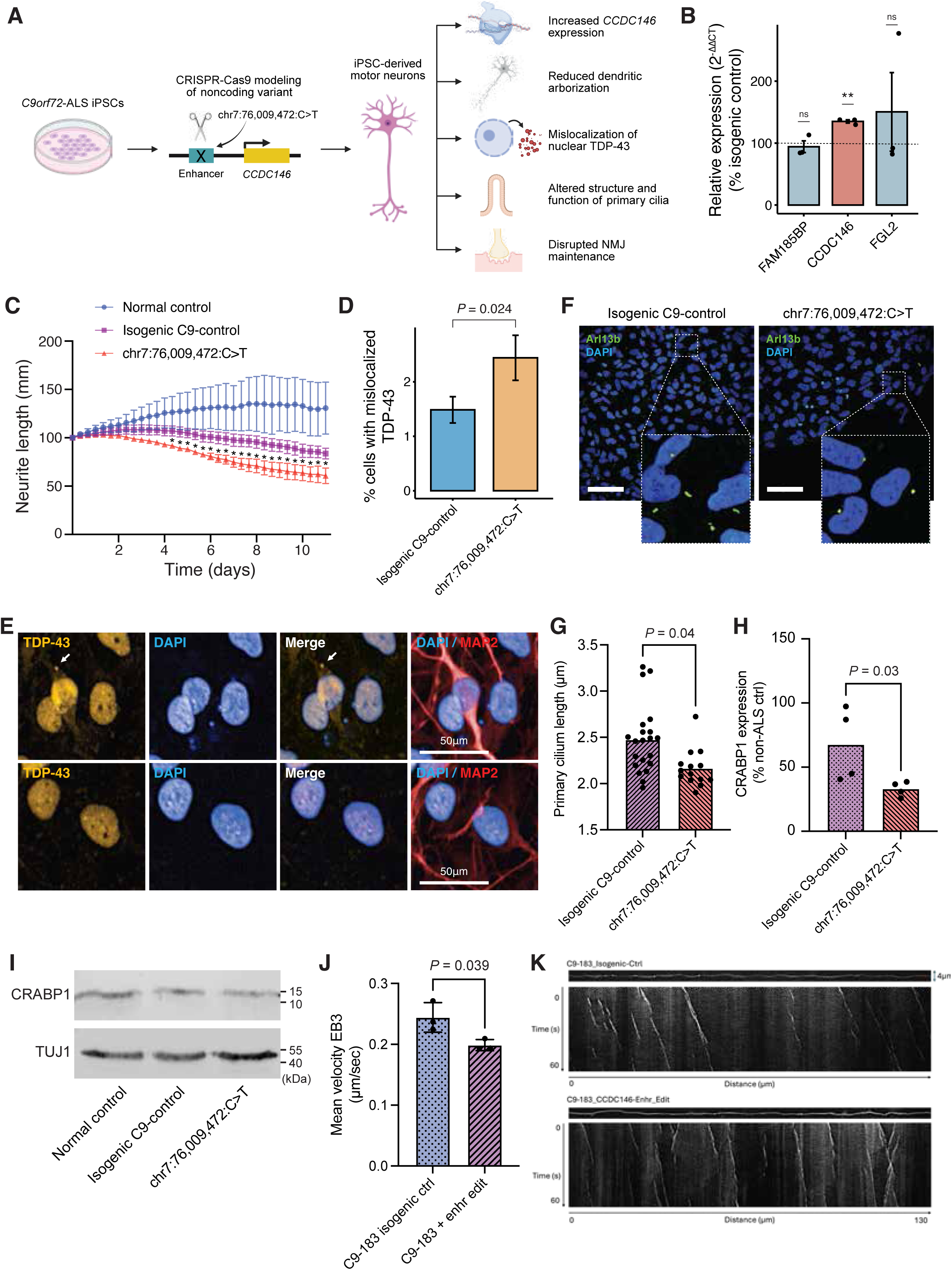
Experimental modeling of chr7:76,009,472:C>T in *C9orf72*-ALS iPSC-derived motor neurons. (**A**) Schematic of CRISPR/Cas9-mediated modeling of chr7:76,009,472:C>T in *C9orf72*-ALS iPSC-derived motor neurons (iMNs). (**B**) Gene expression levels measured by q-RT-PCR for three genes linked to the enhancer harboring chr7:76,009,472:C>T (*n* = 3 technical replicates). The bar plot and error bar represent the mean and standard error, respectively. *P*-value by two-sided *t*-test. ns, not significant; **, *P* < 0.01. (**C**) Quantification of neurite length (*n* = 3 technical replicates) in an age-and sex-matched normal control, an isogenic control carrying a G4C2-repeat expansion of *C9orf72*, and edited iMNs, across 12 days of tracking using the Incucyte live-cell analysis system (Sartorius). The dot and error bar represent the mean and standard error, respectively. *P*-value by two-sided *t*-test. *, *P* < 0.05. (**D**) Quantification of the proportion of MNs exhibiting mislocalization of nuclear TDP-43 to the cytoplasm in isogenic control and edited iMNs. The bar plot and error bar represent the mean and standard error, respectively. *P*-value by two-sided *t*-test. (**E**) Representative images of TDP-43 staining (yellow) in edited iMNs; nucleus (DAPI, blue) and cytoplasm (MAP2, red) staining are also shown. MNs with cytoplasmic mislocalization of TDP-43 are shown (top panels), together with a cytoplasmic inclusion (arrow); for comparison MNs are shown where TDP-43 is localized to the nucleus (bottom panels). Scale bar 50μM. (**F**) Representative images of iMNs where primary cilia were demarcated with Arl13b. Scale bar: original images 100μm; magnified inserts 25μm. (**G**) Measurement of length of primary cilia in iMNs carrying chr7:76,009,472:C>T compared to an isogenic control carrying a *C9orf72* expansion (each dot is the mean length from a single field of view, *n* = 3 separate motor neuron differentiations). *P*-value by two-sided paired *t*-test. (**H**) Measurement of CRABP1 expression in iMNs carrying chr7:76,009,472:C>T compared to an isogenic control carrying a *C9orf72* expansion (*n* = 4 technical replicates). *P*-value by two-sided paired *t*-test. (**I**) Immunoblotting for CRABP1 across different conditions, with TUJ1 for normalization. (**J**) Mean velocity of EB3 comets (μm/sec) in iMN carrying a *C9orf72* expansion with or without chr7:76,009,472:C>T (*n* = 3 technical replicates; ≥4 axons imaged per condition per replicate). The bar plot and error bar represent the mean and standard error, respectively. *P*-value by two-sided *t*-test. (**K**) Straightened axon and representative kymographs depicting distance moved (µm) by EB3 comets over time (seconds) in iMN carrying a *C9orf72* expansion with or without chr7:76,009,472:C>T.

We first examined whether chr7:76,009,472:C>T altered the expression of candidate target genes implicated by predicted E-P links. To achieve this, we differentiated edited iPSCs carrying chr7:76,009,472:C>T, as well as isogenic control iPSCs, into mature MNs using our published small molecule protocol^21^ (**Supplementary Fig. 4A**; **Methods**). The ABC model predicted an E-P link for four genes: *CCDC146, FAM185BP, FGL2,* and *UPK3B*. Study of human tissue revealed that UPK3B is almost exclusively expressed in urothelium^35^, and therefore we excluded this gene from further study. We extracted RNA from mature iMNs and measured the expression levels for *CCDC146, FAM185BP,* and *FGL2* (**Methods**). Expression of *CCDC146* was significantly elevated in the presence of chr7:76,009,472:C>T compared to isogenic control iMNs (*P* = 5.6 x 10^-3^, fold change [FC] = 1.35, two-sided *t*-test; **Fig. 4B**), but *FAM185BP,* and *FGL2* were not differentially expressed.

Next, we sought to determine the effect of chr7:76,009,472:C>T on ALS-associated phenotypes within iMNs. Our genetic analysis suggested that chr7:76,009,472:C>T increased the rate of ALS progression, resulting in a more severe phenotype; therefore we hypothesized that chr7:76,009,472:C>T would increase the severity of ALS-associated phenotypes in iMNs compared to isogenic control iMNs. For an additional comparison, we also included iMNs from an age-and sex-matched neurologically normal individual.

ALS is associated with a number of MN molecular phenotypes, including increased cell death^36^, reduced neurite outgrowth^37, 38^, and TDP-43 mislocalization^39^. We took advantage of this battery of phenotypes to assess the effect of chr7:76,009,472:C>T on ALS severity *in vitro*. iMNs were placed in culture and monitored continuously for 12 days (**Methods**). We measured cell survival and neurite growth at 8 hour intervals. Notably, compared to isogenic control iMNs, the presence of chr7:76,009,472:C>T was associated with a reduction in both neurite length (*P* < 0.05, two-sided *t*-test; **Fig. 4C**) and the number of neurite branch points (**Supplementary Fig. 4B**), and with increased neuron death (**Supplementary Fig. 4C**). iMNs carrying chr7:76,009,472:C>T had a significantly higher burden of TDP-43 mislocalization at the end stage (*P* = 0.024, two-sided *t*-test; **Fig. 4D** and **4E**; **Methods**), compared to isogenic control iMNs.

Previous literature suggests that CCDC146 is located within the basal body of primary cilia^18^. We confirmed that this is also the case within iMNs (**Supplementary Fig. 4D**; **Methods**). Moreover, in iMNs that carried chr7:76,009,472:C>T with elevated *CCDC146* expression, the length of primary cilia was reduced compared to the isogenic control iMNs (*P* = 0.04, paired *t*-test; **Fig. 4F** and **4G**; **Methods**). Primary cilia function is related to CRABP1 expression and hence the maintenance of neuromuscular junctions (NMJs) between MNs and their target muscles^40^. NMJ loss is a key phenotype of ALS^41^ and reduced CRABP1 has been linked to ALS^19^. Immunoblotting for CRABP1 revealed that in iMNs carrying chr7:76,009,472:C>T, CRABP1 expression was reduced compared to the isogenic control iMNs (*P* = 0.03, paired *t*-test; **Fig 4H** and **4I**; **Methods**). Interestingly, ALS-associated Q331K-TARDBP mutations, which disrupt function of TDP-43, have been associated with large downregulation of CRABP1 mRNA^42^ suggesting that the link between ciliary function and CRABP1 expression may be mediated via TDP-43 dysfunction.

We have shown that CCDC146 is located within the basal body of primary cilia and that *CCDC146* levels have an effect on cilia structure and function. CCDC146 is a microtubule-associated protein and cilia are formed from microtubules. Based on this, we further examined microtubule dynamics. EB3 is a well-established regulator of primary cilium biogenesis, facilitating the anchoring of microtubule minus-ends at the basal body and promoting vesicular trafficking to the ciliary base^43, 44^; EB3 depletion has been shown to reduce primary cilium length^43^. Interestingly, we identified a significant reduction in EB3 velocity in chr7:76,009,472:C>T-iMNs with increased *CCDC146* expression (FC = 0.8, *P* = 0.039, unpaired *t*-test; **Fig. 4J** and **4K**; **Methods**), suggesting that the disrupted microtubule dynamics is the link between *CCDC146* and cilia structure and function.

### ASO suppression of CCDC146 corrects cilia dysfunction and rescues ALS-associated survival deficits in patient-derived neurons

We have linked chr7:76,009,472:C>T to elevated expression of *CCDC146* and to ALS-associated molecular phenotypes in iMNs, including TDP-43 mislocalization and cilia dysfunction. To further test whether the expression change of *CCDC146* is the key driver of this toxicity, we synthesized an antisense oligonucleotide (ASO) targeting *CCDC146* mRNA for degradation.

We have established a survival readout for iPSC-derived neurons (iNs) whereby neurons derived from patients display an increased sensitivity to glutamate-induced excitotoxicity^36, 45^ (**Fig. 5A**; **Methods**). This reflects long standing observations regarding glutamate toxicity in ALS^46–48^. We have also established this readout in a 3D spheroid format (3D-iN) to enable prolonged culturing without primary glial cells^45, 48^ (**Fig. 5A**; **Methods**). In our experiment, we treated 3D-iNs derived from multiple patients, including four healthy controls, four ALS patients carrying an *C9orf72* G4C2-repeat expansion, two ALS patients carrying ALS-associated mutations within TARDBP, and five sporadic ALS (sALS) patients (**Supplementary Table 6**). 3D-iNs were exposed to either *CCDC146* ASO or a scrambled control ASO, followed by longitudinal tracking (**Fig. 5A**; **Methods**). First, we confirmed the reduction in *CCDC146* mRNA level within 3D-iNs treated with the *CCDC146* ASO compared to the scrambled ASO (36.9% reduction, *P* = 0.031, two-sided *t*-test; **Fig. 5B**; **Methods**). Notably, we observed that the survival defect was completely rescued by *CCDC146* suppression compared to the scrambled ASO (*P* < 0.05, two-sided *t*-test; **Fig. 5C-E** and **Supplementary Fig. 5A**). The result persisted across all of the different genetic backgrounds tested in our experiment (11 patient lines in total; **Fig. 5C-D** and **Supplementary Fig. 5A**). Moreover, there was no evidence of toxicity mediated by the *CCDC146* ASO in control 3D-iNs (*P* > 0.05, two-sided *t*-test; **Fig. 5D** and **Supplementary Fig. 5A** and **5B**). Indeed, loss-of-function (LoF) of CCDC146 has been shown to be well tolerated in humans^18^. Of note, none of the experimentally-tested patient-derived 3D-iN lines exhibited elevated *CCDC146* expression at baseline (**Supplementary Fig. 5C**). Together, these results demonstrate that *CCDC146* suppression may be a broadly applicable therapeutic strategy for different forms of ALS.

**Fig. 5.**
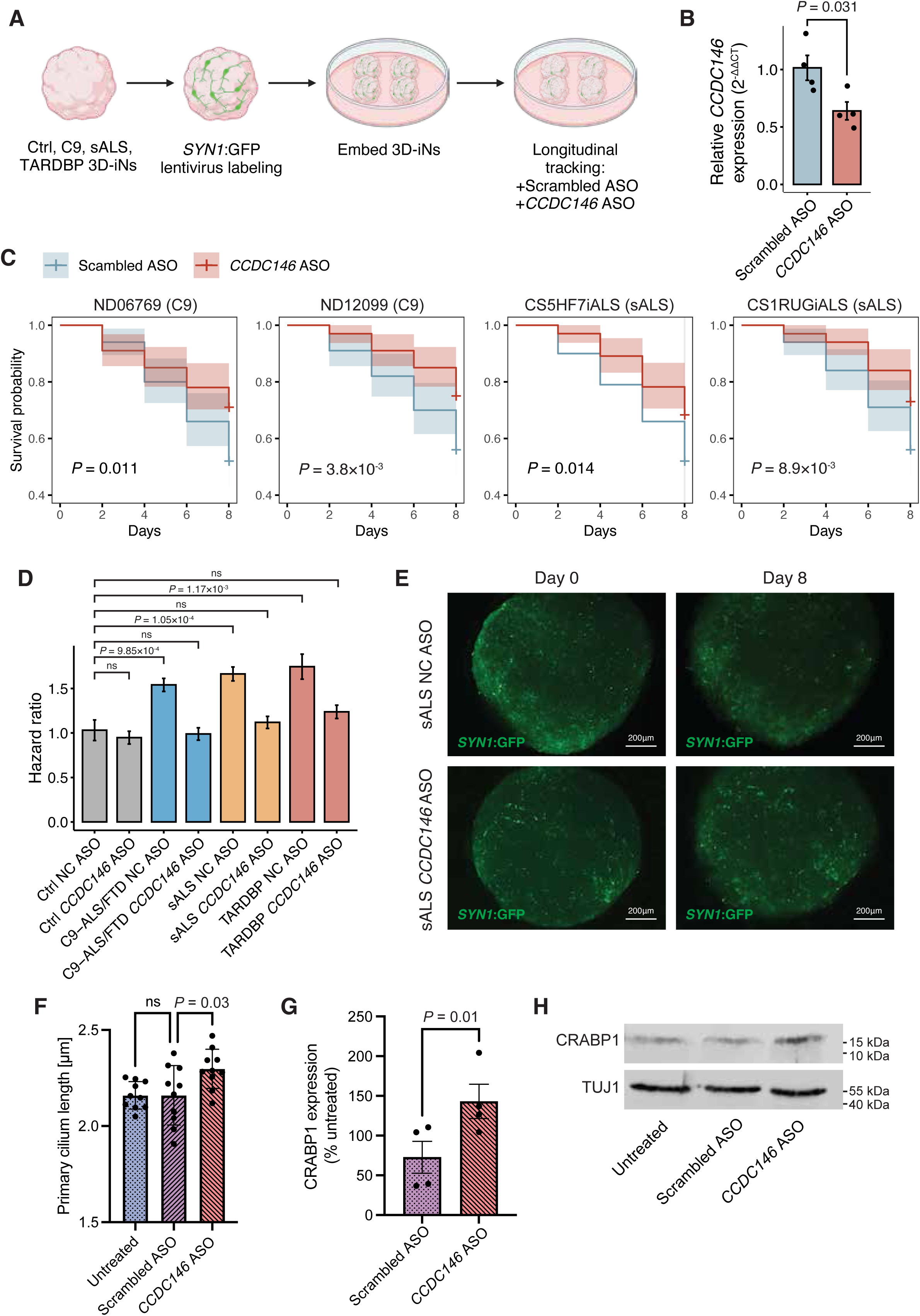
Suppression of *CCDC146* in ALS patient-derived neurons via antisense oligonucleotide (ASO). (**A**) Schematic of our modeling strategy which utilized patient-derived GFP-labeled iPSC-derived neurons in 3D spheroids (3D-iNs). Longitudinal tracking enables accurate cell counting to determine survival following a glutamate insult together with a scrambled ASO or a *CCDC146*-targeting ASO treatment. (**B**) Comparison of gene expression levels measured by q-RT-PCR between *CCDC146*-targeting ASO and scrambled ASO treatments. The bar plot and error bar represent the mean and standard error, respectively. *P-*value by two-sided *t*-test. *n* = 4 technical replicates per condition. (**C**) Kaplan-Meier (KM) estimator plots for four patient cell lines. The shaded area represents 95% confidence interval (CI). *P*-value by log-rank test. C9, *C9orf72*; sALS, sporadic ALS. (**D**) Comparison of hazard ratio measuring the incidence of neuron loss among different conditions and treatments. The bar plot and error bar indicate the mean and standard error, respectively. NC, negative control (scrambled); ctrl, control; C9, *C9orf72*; sALS, sporadic ALS. *P-*value by two-sided *t*-test. ns, not significant. (**E**) Representative images of 3D-iNs derived from a sporadic ALS patient treated with the scrambled ASO (top panels) or the *CCDC146*-targeting ASO (bottom panels). Images were taken from approximately the same location on the plat at Day 0 (left panels) and Day 8 (right panels), demonstrating neuronal loss which was rescued by *CCDC146* suppression. Scale bar: 200μm. (**F-G**) Measurement of length of primary cilia (**F**) and CRABP1 expression (**G**) in iMNs after treatment with *CCDC146* ASO compared to a scrambled ASO or untreated iMNs. The bar plot and error bar represent the mean and standard error, respectively. *P-*value by two-sided *t*-test. ns, not significant. (**H**) Immunoblotting for CRABP1 across different treatments, with TUJ1 for normalization.

Next, we sought to determine whether ASO suppression of *CCDC146* reverses deficits in primary cilia structure and function (**Fig. 4G** and **4H**) which were associated with chr7:76,009,472:C>T and increased expression of *CCDC146*. We treated iMNs derived from a patient carrying a *C9orf72* G4C2-repeat expansion with *CCDC146* ASO and measured primary cilia length and CRABP1 expression after 72 hours (**Methods**). Cilia length was significantly increased after *CCDC146* ASO treatment compared to untreated iMNs and to treatment with a scrambled ASO (*P =* 0.03, one-way ANOVA; **Fig. 5F** and **Supplementary Fig. 5D**). Similarly, CRABP1 expression was increased after treatment with *CCDC146* ASO compared to scrambled ASO (*P =* 0.01, paired *t*-test; **Fig. 5G** and **5H**).

We conclude that *CCDC146* suppression rescues ALS-associated survival defects *in vitro* and reverses deficits in cilia structure and function, suggesting that ciliary function is the mechanism by which *CCDC146* modifies neuronal survival.

### ASO suppression of murine Ccdc146 rescues motor phenotypes and survival defects associated with TDP-43 mislocalization

Depletion of human *CCDC146* corrected survival defects in ALS patient-derived neurons irrespective of genetic background and including sporadic ALS. The majority of ALS is associated with TDP-43 mislocalization^41^, and therefore we hypothesised that *CCDC146* depletion may be effective in an *in vivo* model of ALS caused by TDP-43 mislocalization. Moreover, in iMNs the elevated *CCDC146* expression was associated with an increase in TDP-43 mislocalization, suggesting that reducing *CCDC146* expression may reverse this aberration. To test these hypotheses, we utilized an aggressive juvenile mouse model (TAR4/4 mouse) of ALS caused by overexpression of human TDP-43 which manifests with mislocalization of human and murine tdp-43^17^ (**Fig. 6A**). This model has been previously used to develop therapeutic strategies for ALS^49^.

**Fig. 6.**
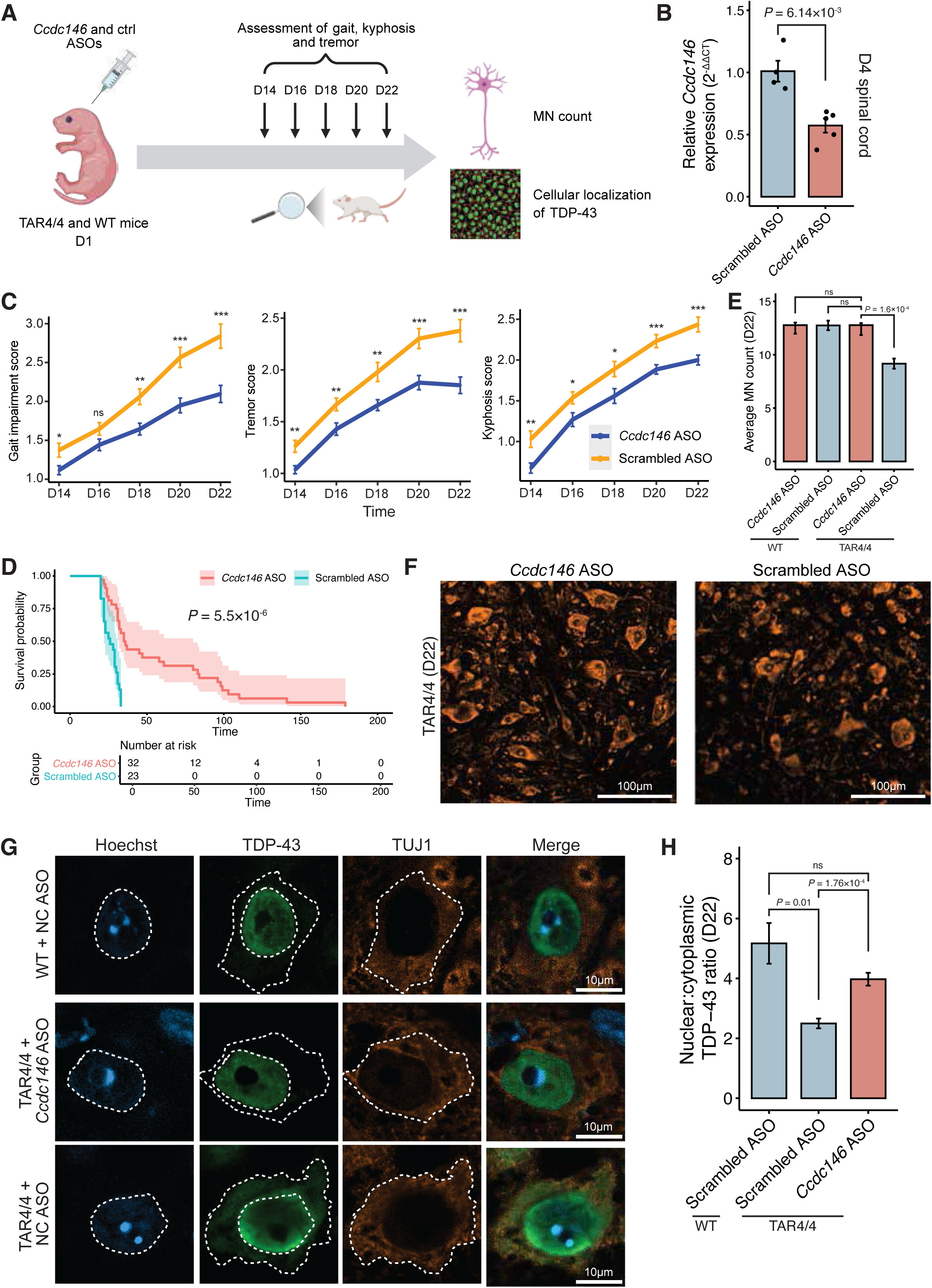
ASO suppression of *CCDC146 in vivo.* (**A**) Strategy for evaluating the *Ccdc146* ASO in Thy1::hTDP43Tg/Tg mice^17^. Intracerebroventricular (ICV) injections of the ASO were performed at postnatal day 1 (D1), with motor phenotype evaluation every two days from postnatal day 14 (D14). WT, wild type; MN, motor neuron. (**B**) Mouse endogenous *Ccdc146* gene expression levels measured after treatment with the *Ccdc146* ASO or a scrambled control ASO at D4 (*n* = 4 technical replicates). The bar plot and error bar represent the mean and standard error, respectively. *P*-value by two-sided *t*-test. (**C**) Evaluations of gait impairment, tremor, and kyphosis after treatment with the *Ccdc146* ASO or a scrambled ASO. The line plot and error bar indicate the mean and standard error, respectively. *P*-value by two-sided *t*-test. *, <0.05, **, <0.01, ***, <0.001. (**D**) Kaplan-Meier (KM) estimator plot for mouse survival after treatment with either *Ccdc146* ASO (*n* = 32) or a scrambled control ASO (*n* = 23). The shaded area represents 95% confidence interval (CI). *P*-value by Cox regression. (**E**) Motor neuron counts in TAR4/4 and wild type mice treated with either *Ccdc146* ASO or a scrambled control ASO. Mice were sacrificed at D22 for histology. The bar plot and error bar represent the mean and standard error, respectively. *P*-value by two-sided *t*-test. WT, wild type; ns, not significant. (**F**) Immunostaining of large TUJ1+ neurons in the ventral horn of TAR4/4 mice treated with a scrambled ASO or *Ccdc146* ASO used to perform motor neuron counts. (**G**) Immunostaining of total TDP-43 in large TUJ1+ neurons in the ventral horn of wild-type and TAR4/4 mice treated with a scrambled ASO or *Ccdc146* ASO. Dotta lines to outline the cell body/nucleus used for quantification. WT, wild type; NC, negative control (scrambled); TAR4/4, Thy1::hTDP43Tg/Tg. (**H**) Nuclear:cytoplasmic TDP-43 ratio at D22 across different treatments. Each data point represents the average TDP-43 nuclear:cytoplasmic ratio for one mouse. The bar plot and error bar indicate the mean and standard error, respectively. *P*-value by two-sided *t*-test. WT, wild type; ns, not significant.

We generated an ASO against murine *Ccdc146* which successfully depleted *Ccdc146* expression when injected intracerebroventricular (IVC) at D1 (*P* = 6.14 x 10^-3^, two-sided *t*-test; **Fig. 6B**; **Methods**). Notably, the *Ccdc146* ASO mitigated motor phenotypes compared to a scrambled ASO and the difference increased over time (**Fig. 6C**); at D22 the differences were significant for all measures (*n* > 30 in each group; gait impairment score: *P* = 2.7 x 10^-4^; tremor score: *P* = 2.2 x 10^-4^; kyphosis score: *P* = 2.1 x 10^-4^, two-sided *t*-test; **Fig. 6C**; **Methods**). Moreover, survival was significantly prolonged in mice treated with the *Ccdc146* ASO (*n* = 55, 36% increase in median survival time, *P* = 5.5 x 10^-6^, Cox regression; **Fig. 6D**; **Methods**), compared to mice treated with the scrambled ASO. To determine whether the *Ccdc146* ASO acts via reversal of MN toxicity, we performed MN counts in wild type and TAR4/4 mice treated with *Ccdc146* or scrambled ASO. Treatment with *Ccdc146* ASO significantly increased the average number of MNs at D22 (*P* = 1.6 x 10^-4^, two-sided *t*-test; *n* > 10 biological replicates, *n* = 6 fields-of-view (FOV) per mouse; **Fig. 6E** and **6F**). Indeed at D22, MN counts in *Ccdc146* ASO-treated TAR4/4 mice were equivalent to wild type animals (**Fig. 6E**). Finally, we sought to examine the distribution of murine TDP-43 within MNs using immunohistochemistry (**Methods**). The *Ccdc146* ASO significantly increased the nuclear:cytoplasmic ratio of murine TDP-43 within MNs at D22 compared to the scrambled ASO (*P* = 1.76 x 10^-4^, two-sided *t*-test; **Fig. 6G** and **6H**), exhibiting an almost complete rescue of TDP-43 mislocalization when compared to the wild type mice (*P* > 0.05, two-sided *t*-test; **Fig. 6G** and **6H**).

We conclude that ASO suppression of *CCDC146* is an effective therapeutic strategy *in vivo,* and that this activity is upstream of TDP-43 mislocalization which is the defining pathology of ALS^39^.

## Discussion

We have designed ASOs targeting *CCDC146* which ameliorate ALS-associated phenotypes, including survival and TDP-43 mislocalization both *in vitro* and *in vivo*. We achieved this by starting with a hypothesis-free *in-silico* screen for genetic modifiers of ALS survival. Distinct from previous works, our genetic screen was built upon a deep learning-based variant interpretation framework that makes MN-specific functional effect predictions for an exhaustive set of noncoding variants. Our model, named NeuroNet, was trained to predict various MN epigenomic features (e.g., chromatin accessibility and histone modification) from DNA sequence. This enables the model to make functional predictions for unseen genetic variants irrespective of allele frequency. Deep learning-based seq2func modeling has been widely adopted to examine the cell-type-specificity of regulatory genome function and genome variation^11–16, 22, 50^, and here we demonstrated its success in identifying potential therapeutic targets for ALS. Conventional approaches for investigating variant effects, such as the expression and chromatin accessibility quantitative trait loci (eQTL and caQTL) analyses, require matched individual genotype and transcriptomic/epigenomic profiling data, which is typically underpowered for the study of rare or ultra-rare variants.

Combining NeuroNet predictions of variant effects on the epigenome, together with enhancer-promoter links, enabled us to build a variant-to-gene (V2G) map within MNs. The result is a gene-resolved prediction of noncoding variant function. Based on this V2G, we have conducted large-scale rare variant burden tests for both ALS risk and survival. Although the risk analysis was underpowered due to the paucity of control samples, a NeuroNet-enabled survival analysis succeeded whereas a traditional analysis was still underpowered. This demonstrates the benefit of incorporating functional information into a genetic analysis.

We have identified four noncoding variants linked to ALS survival, and they were predicted to modulate the expression of three protein-coding genes: *ZNF839*, *NRXN1*, and *CCDC146*. Interestingly, although we did not identify any genome-wide significant genes associated with ALS risk, we did note a significant overlap between nominally significant risk- and survival-associated genes. This is consistent with known genetic variants which affect both ALS risk and survival including noncoding variants associated with *UNC13A*^51^ and *C9orf72^3^*, and provides evidence for the overlapping genetic architecture of ALS risk and survival within the noncoding genome.

We reproduced the top hit chr7:76,009,472:C>T in iMNs carrying an ALS-associated G4C2 expansion within *C9orf72*, and verfied its regulation of *CCDC146* expression. We showed that chr7:76,009,472:C>T exacerbated ALS-associated phenotypes in iMNs, including reduced MN survival compared to isogenic control neurons. Moreover, using an ASO to reduce *CCDC146* expression completely rescued ALS-associated survival defects in iMNs, irrespective of genetic background and without significant toxicity, and also irrespective of the baseline *CCDC146* expression. Similarly, we observed a dramatic survival benefit in an aggressive mouse model of ALS caused by mislocalization of nuclear TDP-43^17^, which is the key molecular pathology underlying >98% of ALS^39^. We note that a different ASO has recently been approved for treatment of ALS patients carrying a pathogenic mutation within SOD1^52^. Unlike that treatment - Tofersen - our data suggest an ASO targeting *CCDC146* could be administered to broad forms of ALS and not limited to a specific genetic subtype.

CCDC146 is a microtubule associated protein (MAP) and localized to sperm flagella and to primary cilia in particular. Biallelic variants in *CCDC146* have been associated with reduced sperm viability and non-syndromic male infertility in both mice and humans^18^. Flagella are functionally mirrored by cilia and it is notable that primary cilia dyskinesia has been associated with ALS^19, 53, 54^. Here, we have determined that CCDC146 is localized to primary cilia in human MNs, in a similar manner to *C21ORF2/CFAP410*^19^ which is an ALS risk gene^55^ and is important for cilia assembly. Our data suggest that this may be because CCDC146 is a modifier of microtubule dynamics, and cilia are microtubule structures. Primary cilia occur one per cell in neurons and extend into the extracellular space where they can transduce intercellular signals via G-protein coupled receptors. Unlike in sperm cells, neuronal cilia are non-motile structures with an important role in sensing extracellular signals and in transduction of *sonic hedgehog* (shh) signalling. Elevated *CCDC146* causes shortened primary cilia which would be expected to impair intraflagellar transport (IFT)^19^. IFT is the process by which structural components are selected and positioned for the assembly and maintenance of cilia structure; indeed IFT occurs continuously in two directions towards and away from the tip of each cilium and occurs in proportion to cilium length^56^.

We identified loss of CRABP1 in MNs overexpressing *CCDC146* and increase in CRABP1 after *CCDC146* depletion by ASO. CRABP1 is an important protein for NMJ structure and function within MNs. CRABP1 expression is relatively specific to the spinal cord and to MN in particular^40^; indeed CRABP1 knockout mice exhibit an ALS-like phenotype, including MN degeneration^40^. *CRABP1* expression has been linked to ciliary function via shh signalling^57^, and we also note reports in the literature of a link between TDP-43 dysfunction and *CRABP1* expression^42^. Indeed, *CRABP1* is implicated in maintenance of NMJ function via *CAMK2B* and *AGRN*^40^, both of which are misplaced in the context of TDP-43 dysfunction^58^; this convergence supports the idea that this is a key pathway in driving neuronal toxicity in ALS. We suggest that the positive effect of *CCDC146* on MN health is mediated via modification of ciliary function leading to recovery of TDP-43 mislocalisation and elevation of CRABP1 expression. TDP-43 mislocation is a key driver of >98% of ALS^39, 58, 59^ and so our proposed mechanism would explain why the *CCDC146* ASO appears to be broadly applicable to patient-derived neurons. Here we have provided evidence to link *CCDC146* expression to TDP-43 mislocalization both *in vitro* and *in vivo*. While further research is needed to confirm and clarify the interaction between ciliary function and TDP-43 mislocalization, our work opens a new avenue for understanding the role of primary cilia as a universal driver of MN death in ALS.

It is notable that, aside from a nonsyndromic fertility defect, loss of CCDC146 is well tolerated^18^; this crucial observation suggests that our ASO approach may be viable without significant toxicity. Additionally, this gene is not well conserved^60^ (probability of loss of function intolerance (pLI) = 0). CCDC146 is expressed throughout the brain and spinal cord, and particularly within neurons^61^ (Human Protein Atlas). Expression of *CCDC146* has been identified specifically in neurons from the human spinal cord^62^. Matching our finding of higher *CCDC146* expression associated with reduced ALS survival, the expression of *CCDC146* is higher in ALS patient spinal cord compared to control cervical spinal cord although, at a bulk level, this difference was not statistically significant^63^.

We investigated the potential of targeting CCDC146 through small-molecule drugs. We first searched chemical and patent databases but found no active compounds targeting CCDC146 and similar proteins (**Methods**). CCDC146 is also classified as a poorly explored target without active small molecules according to the Pharos database^64, 65^. Due to the lack of experimentally solved protein structure data, we obtained the AlphaFold-predicted structure of CCDC146^66^ (**Supplementary Fig. 6**). The predicted structure mainly consists of coiled coils linked by flexible regions, without ligand binding sites detected on its surface^67^ (**Methods**), suggesting that it would be challenging to design small-molecule drugs for this target and underscoring the importance of alternative targeting approaches such as an ASO.

We also sought to validate another gene *ZNF839* in the context of disease progression. In particular, we inspected gene expression in iMNs derived from an independent cohort encompassing 245 ALS patients (www.answerals.org; **Methods**). We measured disease progression using two metrics, including the survival time from symptom onset and the rate of change in ALS Functional Rating Scale in its revised version (ALSFRS-R)^68^. ALSFRS-R is widely used to measure ALS progression^69^. We found that lower expression of *ZNF839* was associated with faster rate of disease progression (*β* = 0.85, *P* = 5.9 x 10^-3^, multivariable linear regression; **Supplementary Fig. 7A**; **Methods**) and shorter survival time (*β* = -0.79, *P* = 0.044, Cox PH regression; **Supplementary Fig 7B**; **Methods**). The expression levels of other genes we identified were not correlated with either the rate of change in the ALSFRS-R or survival, probably because the AnswerALS-based analysis was underpowered.

Given the significant enrichment of European samples in our WGS cohort, a potential limitation of our work is that our conclusions might not be generalizable to other ancestral groups. However, the allele frequencies of all four survival variants are below 1% (the maximum population-level allele frequency is 5.9% obtained for chr14:102,782,854:G>A) across different populations according to the gnomAD whole-genome collection 2.1.1, implicating the ancestral transferability of the effect of our identified variants, although further confirmation is required.

In summary, we have developed a deep learning-powered genetic analysis framework for systematic identification of cell-type-resolved genetic modifiers of ALS survival. Each of the candidate genes we pinpointed represents a potential therapeutic target, but we have demonstrated *in vitro* and *in vivo* that ASO knockdown of *CCDC146* is a safe, broadly applicable and efficacious therapeutic strategy. NeuroNet can be applied to study the genetics of other MN-specific diseases, including spinal muscular atrophy (SMA) and hereditary sensory and motor neuropathy (HSMN). Our analysis framework contributes to a better understanding of ALS genetic architecture, defines a novel therapeutic target for ALS, and provides a generic paradigm for enhancing noncoding genetic analysis of complex human diseases.

## Supporting information

Supplementary Results

## Data Availability

The epigenome and transcriptome profiling data of iPSC-derived motor neurons used in the ABC model and NeuroNet are available at encodeproject.org with the following link: https://www.encodeproject.org/publications/de19555b-a49f-471c-bfbc-be3b628fe9bf/. NeuroNet variant effect scores can be downloaded at https://doi.org/10.6084/m9.figshare.25444534. The Project MinE WGS data are accessible upon application approval (https://www.projectmine.com/research/data-sharing/). The AnswerALS WGS and RNA-seq data are available upon request approval (https://dataportal.answerals.org/home).

https://doi.org/10.6084/m9.figshare.25444534

https://www.projectmine.com/research/data-sharing/

https://dataportal.answerals.org/home

## Acknowledgments

This work was supported by NIH (CEGS 5P50HG007735 to M.P.S., R01NS131409 and R01NS097850 to J.K.I.), Wellcome Trust (216596/Z/19/Z to J.C.-K.), NIHR (NF-SI-0617-10077 to P.J.S.), Motor Neurone Disease (MND) Association (894-791, 899-792 and 913-792 to C.H., S.G., S.Z. and J.C.-K.), John Douglas French Alzheimer’s Foundation (to J.K.I.), and Robert Packard Center for ALS Research (1375407 to S.Z.). We also acknowledge support from the MND Association Fellowship (2323-799) and the Kingsland Fellowship (to T.M.), the NIHR Sheffield Biomedical Research Centre (NIHR203321), and the NIHR Sheffield Clinical Research Facility. We are very grateful to the ALS patients and control subjects who generously donated biosamples. We acknowledge transcriptomic and clinical data provided by the AnswerALS Consortium. Figures 1, 2a, 4A, 5A and 6A were created using BioRender.com.

## Author contributions

S.Z., J.C.-K. and M.P.S. conceived and designed the study. S.Z. and E.Y. developed NeuroNet. S.Z., J.C.-K., T.M. and J. R.-S. performed statistical analyses. T.M. performed all the experiments for reproducing and characterizing chr7:76,009,472:C>T in iMNs. J. R.-S., S.T. and T.M. carried out ASO-related experiments in 3D-iNs, iMNs and mice. S.Z., J.C.-K., T.M. and J. R.-S. interpreted the data with assistance from all other authors. The Project MinE ALS Sequencing Consortium was involved in WGS data acquisition and analysis. S.Z., J.C.- K. and M.P.S. supervised the work. S.Z., J.C.-K. and M.P.S. wrote the manuscript with feedback from all other authors.

## KEY RESOURCES TABLE

| Reagent or Resource | Source | Identifier |
| --- | --- | --- |
| *Antibodies* |  |  |
| anti-Nestin | Abcam | #ab18102 |
| anti-Pax6 | Abcam | #ab5790 |
| anti-Beta III Tubulin | Abcam | #ab18207 |
| anti-HB9 | Abcam | #ab221884 |
| anti-Islet 1/2 | Abcam | #AB109517 |
| anti-NeuN | Millipore | #MAB377 |
| anti-ChAT | Millipore | #AB144P |
| anti-SMI-32 | Biolegend | #801701 |
| anti-phospho TDP-43 (Ser409) | Affinity Biosciences | #AF7365 |
| anti-TDP-43 | Proteintech | #12892-1-AP |
| anti-MAP2 | Proteintech | #17490-1-AP |
| anti-GFP | Novus Biologicals | #AB_10128178 |
| anti-NFH | Millipore Sigma | #AB_477272 |
| anti-CCDC146 | Sigma | #HPA020082 |
| CoraLite® Plus 488-conjugated anti-ARL13B | Proteintech | #CL488-17711 |
| Donkey anti-Mouse IgG Alexa Fluor 488 | Thermofisher | #A-21202 |
| Donkey anti-Rabbit IgG Alexa Fluor 568 | Thermofisher | #A-10042 |
| Donkey anti-Goat IgG Alexa Fluor 555 | Thermofisher | #A-21432 |
| Goat anti-Guinea Pig IgG Alexa Fluor 647 | Thermofisher | #A-21450 |
| Donkey anti-Mouse IgG Alexa Fluor 594 | Thermofisher | #A-32744 |
| Donkey anti-Rabbit IgG, Alexa Fluor 488 | Thermofisher | #A-21206 |
| Donkey anti-Mouse IgG Alexa Fluor 568 | Thermofisher | #A-10037 |
| Goat anti-Chicken IgY (H+L) Alexa Fluor Plus 488 | Thermofisher | #A-32931 |
| Alexa Fluor 680-AffiniPure Donkey Anti-Rabbit IgG (H+L) | Jackson ImmunoResearch | #711-625-152 |
| *Chemicals, Peptides and Recombinant Proteins* |  |  |
| Hoechst 33342 | ThermoFisher Scientific | #62249 |
| Triton X-100 | Sigma-Aldrich | #T8787 |
| Donkey serum | Millipore | #S30-100ML |
| Bovine serum albumin | Sigma | #A2153 |
| KnockOut™ DMEM/F-12 | Gibco | #12660012 |
| Neurobasal medium | Gibco | #21103049 |
| Penicillin-Streptomycin | Sigma | #P4333 |
| Gibco GlutaMAX™ Supplement | ThermoFisher Scientific | #35050061 |
| N-2 supplement | ThermoFisher Scientific | #17502048 |
| B-27 supplement | ThermoFisher Scientific | #17504001 |
| mTeSR-Plus Medium | StemCell Technologies | #100-0276 |
| Accutase™ Cell Dissociation Reagent | Gibco | #11599686 |
| ReLeSR™ | StemCell Technologies | #100-0483 |
| Matrigel® | Corning | #356230 |
| Poly-L-ornithine | Sigma | #P4957 |
| Purmorphamine | Tocris Bioscience | #4551 |
| DMH-1 | Tocris Bioscience | #4126 |
| SB431542 | Tocris Bioscience | #1614 |
| CHIR99021 | Tocris Bioscience | #4423 |
| ROCK inhibitor (Y-27632 dihydrochloride) | Tocris Bioscience | #1254 |
| Compound E | Tocris Bioscience | #6476 |
| All-trans retinoic acid | Sigma | #R2625 |
| Valproic acid | Sigma | #P4543 |
| hBDNF | R&D systems | #248-BDB-010/CF |
| IGF-1 | R&D systems | #291-G1 |
| CNTF | R&D systems | #257-NT-010/CF |
| Dulbecco's Phosphate Buffered Saline | Sigma | #D8537-500ML |
| EDTA | Sigma | #E5134 |
| HEPES | Sigma | #H3375 |
| PMSF protease inhibitor | ThermoFisher Scientific | #36978 |
| Protease inhibitor tablet | Roche | #1697498 |
| TEMED | ThermoFisher Scientific | #17919 |
| Ammonium Persulfate | ThermoFisher Scientific | #17874 |
| 30% Acrylamide/Bis-acrylamide solution | Sigma | #A3699-100ML |
| TE Buffer, RNase-free pH 8 | ThermoFisher Scientific | #AM9849 |
| Ethidium bromide solution | Sigma | #E1510 |
| VeriFi mix red | PCRBio | #PB10.42-01 |
| Tri reagent | Sigma | #93289-100ML |
| M-MLV reverse transcriptase | ThermoFisher Scientific | #28025-013 |
| 5x First Strand buffer | ThermoFisher Scientific | #18057-018 |
| 0.1M Dithiothreitol | ThermoFisher Scientific | #707265ML |
| dNTP Mix | ThermoFisher Scientific | #10534823 |
| Random hexamer primer | ThermoFisher Scientific | #SO142 |
| SYBR Green Brilliant III master mix | Agilent | #600882 |
| *Critical Commercial Assays* |  |  |
| Protein Bradford Assay | Bio-Rad | #5000201 |
| GenElute Mammalian Genomic DNA Miniprep Kit | Sigma | #G1N350 |
| Direct-zol RNA Miniprep Kit | Zymo Research | #R2050 |
| QIAquick PCR Purification kit | Qiagen | #28104 |
| QIAquick Gel Extraction Kit | Qiagen | #28506 |
| *Experimental Models: Cell Lines* |  |  |
| GM23338 (iPSC line derived from healthy volunteer) | Coriell Institute | #CVCL_F182 |
| KOLF2.1J (iPSC line derived from healthy volunteer) | The Jackson Laboratory | #HPSI0114i-kolf_2 |
| C9-183 (iPSC line derived from a patient with C9orf72-ALS) | The University of Sheffield | #RRID:CVCL_UF85 |
| C9-183_chr7:76,009,472:C>T (iPSC line derived from a patient with C9orf72-ALS with CRISPR-engineered enhancer edit [chr7:76,009,472:C>T]) | The University of Sheffield | - |
| *Software and Algorithms* |  |  |
| R v4.0.1 | https://cran.r-project.org/mirrors.html |  |
| PRISM 10 | GraphPad |  |
| ICE CRISPR analysis tool | https://ice.synthego.com/#/ |  |
| CFX Maestro | Bio-Rad |  |
| Harmony Imaging Analysis Software | PerkinElmer (Revvity) |  |
| FIJI (FIJI Is Just ImageJ) | NIH |  |
| Incucyte® software | Sartorius |  |
| Image Studio™ Software | LI-COR |  |
| IGV v2.4.16 | https://software.broadinstitute.org/software/igv/ |  |
| NeuroNet | https://github.com/szhang112/neuronet |  |

## RESOURCE AVAILABILITY

### Lead contact

Further information and requests for resources and reagents should be directed to and will be fulfilled by the Lead Contact, Michael P. Snyder.

### Material availability

All unique/stable reagents generated in this study are available from the Lead Contact without restriction.

### Data availability

The epigenome and transcriptome profiling data of iPSC-derived motor neurons (iMN) used in the ABC model and NeuroNet are available at encodeproject.org with the following link: https://www.encodeproject.org/publications/de19555b-a49f-471c-bfbc-be3b628fe9bf/. NeuroNet variant effect scores can be downloaded at https://doi.org/10.6084/m9.figshare.25444534. The Project MinE WGS data are accessible upon application approval (https://www.projectmine.com/research/data-sharing/). The AnswerALS WGS and RNA-seq data are available upon request approval (https://dataportal.answerals.org/home).

### Code availability

The NeuroNet source code and tutorial are available at https://github.com/szhang1112/neuronet.

## METHODS

### Prediction of enhancer-promoter interactions using the ABC model

We implemented the ABC model following the guidelines provided at https://github.com/broadinstitute/ABC-Enhancer-Gene-Prediction. First, we called peaks for the iMN ATAC-seq profiling data^21^ using MACS2, and then identified the candidate enhancer elements using “makeCandidateRegions.py” with peakExtendFromSummit = 250 and nStrongestPeaks = 150000. The black-listed regions generated by the ENCODE 4 (https://www.encodeproject.org/) were used for removing enhancers overlapping regions with anomalous sequencing reads. Second, we applied “run.neighborhoods.py” to quantify the enhancer activities by counting ATAC-seq and H3K27ac ChIP-seq reads in candidate enhancer regions. RNA-seq profiling of iMNs was also provided to inform expressed genes. Quantile normalization was applied using K562 epigenomic data as the reference. At last, using “predict.py” we computed the ABC scores by combining the enhancer activities (calculated by the second step) with the Hi-C profiling. Hi-C data were fit to the power-law model. The suggested threshold 0.02 was used to define valid E-P links.

### NeuroNet model design and training

The architecture of NeuroNet follows the Beluga structure^12^, a powerful convolutional neural network (CNN) designed to predict 2,002 epigenomic features from the DNA sequence, except the last layer of NeuroNet is tailored to predict four epigenomic features (chromatin accessibility, H3K27ac, H3K4me1, and H3K4me3) in iMNs.

The input of NeuroNet is the one-hot encoded 2,000-base-pair (bp) DNA sequence of which the centered 200 bp region overlaps peaks of at least one epigenomic feature – called valid sequence in this paper. The model outputs four probabilities indicating the peak status. Given the encoded input sequence *X_i_* (*i* = 1, …, *N*), the model *f* is trained to minimize the following binary cross-entropy loss:

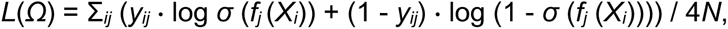

Where *j* is the index of four epigenomic features, *σ* is the sigmoid function, *yij* is the binary label of the *j*-th feature of the *i*-th valid sequence, and *Ω* is the set of network parameters.

Genome-wide valid sequence samples were randomly split into 1270000, 4000, and 164080 training, validation, and testing samples, respectively. The dataset was further augmented by adding reverse complement sequences. Stochastic gradient descent (SGD) with learning rate = 0.1, momentum = 0.9, and weight decay = 1 x 10^-6^ was adopted as the optimizer. Batch size was set to 256. We also applied early stopping to prevent overfitting, in which the AUPRC was selected as the monitoring metric for an imbalanced dataset.

The network structure of NeuroNet follows the Beluga architecture which has been widely used in biological sequence modeling. Following the best practice^11, 12, 50^, we trained NeuroNet based on 1,270,000 training examples and evaluated model prediction using 164,080 sequence-epigenome pairs held-out on chromosomes 8 and 9.

### Variant effect prediction using NeuroNet

After the model was trained, we utilized NeuroNet to infer the functional effects of individual variants on epigenomic features. Given a genetic variant, we construct a variant-centered DNA sequence based on the reference genome, along with its reverse complement sequence. The model predictions for both forward and reverse complement sequences are averaged to yield the variant epigenomic scores *S*_alt,*j*_. Similarly, the epigenomic scores *S*_ref,*j*_ are also calculated. We define the variant functional scores *S*_var,*j*_ per epigenomic feature as the absolute difference between reference and alternative epigenomic scores, i.e., *S*_var,*j*_ = |*S*_ref,*j*_ - *S*_alt,*j*_|, where *j* indicates different epigenomic features. The final NeuroNet score *S*_NN_ for each variant is defined as the maximum of all functional scores, i.e., *S*_NN_ = max_*j*_ *S*_var,*j*_. As justified in the main text, we chose 0.06 as the threshold to define “functional noncoding variants” in MNs, that is, a noncoding variant is kept for downstream analysis if and only if its NeuroNet score satisfies *S*_NN_ > 0.06.

### Transcription factor binding motif analysis

The TF binding motif analysis was performed using GimmeMotifs^70^. The short-range and long-range enhancers were defined as enhancers with the mean absolute E-P distance below 5% and above 95% of the distribution, respectively. The differential motifs between short- and long-range enhancers were identified using the “gimme maelstrom” command. This command combined the results from different methods using the Stouffer’s method, yielding an integrated *Z*-score. A higher *Z*-score means the motif better matches the sequence of the peak region in one condition than the other.

### Network analysis

We downloaded the human PPIs from STRING version 12.0^71^, including 19,622 proteins and 6,857,702 protein interactions. We extracted high-confidence (combined score >700) PPIs for downstream analysis, including 16,185 proteins and 236,000 protein interactions. To eliminate the bias caused by hub proteins^72^, we ran the random walk with restart (RWR) algorithm over the PPI network, wherein the restart probability was set to 0.5, resulting in a smoothed network after preserving the top 5% predicted edges (*N* = 6,243,766). We then applied the Louvain method^73^, a classic community detection algorithm searching for densely connected modules, to decompose the network into different modules. After the algorithm converged, we obtained 1,261 modules with an average size of 13 nodes.

The enrichment of genes of interest within each module was tested using the hypergeometric test. Modules with false discovery rate (FDR) < 0.1 after the BH procedure were reported.

### Gene ontology analysis

The gene ontology (GO) analysis in this work was performed based on clusterProfiler^74^ and enrichR^75^. All enrichment analyses were focused on the biological process and molecular function. The gene enrichment was evaluated based on a two-sided Fisher’s exact test, and only terms with adjusted *P* < 0.1 (after BH correction) were reported. To remove the GO term redundancy in some cases, we applied the “simplify” function provided by clusterProfile.

### Benchmarking of NeuroNet prediction against other variant scores

To benchmark NeuroNet against DeepSEA, the Pearson correlation between NeuroNet prediction for four MN epigenomic features and DeepSEA prediction for matched epigenomic features was calculated. For each MN epigenomic feature, all correlation coefficients were first sorted and evenly divided into highly- and weakly-correlated sets, and then the rank of brain- and neuron-related correlations was examined using a two-sided Fisher’s exact test within these two sets.

SNP2TFBS^24^ was used to predict the variant effect on TF binding affinity. The binding affinities for different TFs were averaged for each studied variant to estimate its overall effect. Given a particular NeuroNet cutoff, the noncoding variants were split into two groups according to their NeuroNet predictions. We compared the SNP2TFBS scores between these two groups of variants and examined the difference using a two-sided *t*-test. The *t*-statistics were reported to indicate the SNP2TFBS score difference between two groups. The final NeuroNet score threshold was determined at the largest difference.

With the determined NeuroNet threshold, the correlation between NeuroNet prediction and other scores was inspected by comparing scores between NeuroNet-positive and NeuroNet-negative variants using a two-sided Fisher’s exact test.

### Whole-genome sequencing data in Project MinE

Project MinE WGS data (Data Freeze 2) consists of 6,715 ALS patients and 2,885 healthy controls recruited at specialized neuromuscular centers in the UK, Belgium, Germany, Ireland, Italy, Spain, Turkey, the United States and the Netherlands^76^. Patients were diagnosed with possible, probable or definite ALS according to the 1994 El-Escorial criteria^77^. All controls were free of neuromuscular diseases and matched for age, sex and geographical location. Sample-level and variant-level quality controls (QCs) have been described previously^76^. Briefly, variants with missingness > 5% were removed, as were variants out of Hardy-Weinberg equilibrium (HWE) in controls (*P* < 1 x 10^-6^) and monomorphic variants. All samples had missingness <10% across all 22 chromosomes and samples with an inbreeding coefficient >3 standard deviations from the mean of the distribution were excluded, as were unexpected related samples. After QCs, 6,025 ALS genomes were retained for downstream analysis.

### ALS survival and risk analysis based on NeuroNet-prioritized rare noncoding variants

After QCs, single nucleotide variants (SNVs) meeting all the following criteria were reserved for the survival analysis: (1) minor allele frequency (estimated based on non-Finnish European samples in the gnomAD whole-genome collection 2.1.1) < 0.5%; (2) minor allele count ≥ 3 within the QCed cohort; (3) residing within MN CREs (promoters and enhancers) defined by the ABC model; (4) NeuroNet score > 0.06. These criteria together defined the “functional rare noncoding variants” in our study. Next, screened variants were collapsed to their target genes based on the ABC E-P links and the number of total mapped variants for each gene was counted (i.e., gene burden). To associate the gene burden to survival, we applied the Cox proportional-hazards model implemented by the “coxph” function in the R “survival” package, with 26 covariates including age, sex, age^2^, sex x age, sequencing platform, *C9orf72* status and the first 20 principal components. Because many genes shared the same group of regulatory variants resulting in duplicate tests, we collapsed those genes into unique sets and carried out the Bonferroni multiple testing correction. Tests with adjusted *P* < 0.05 were reported.

We also performed a case-control analysis by including the control samples in Project MinE. 2,264 controls were reserved for analysis after QCs. Functional rare noncoding variants were identified in the same way as that in the survival analysis, and then included in the case-control study. Gene burdens were calculated similarly. To associate gene burden to the disease status, we applied the SKAT-O method^30^ implemented in the RVTESTS^30^, with 25 covariates including age, sex, age^2^, sex x age, sequencing platform, and the first 20 principal components.

### Benchmarking of survival analysis

The benchmarking survival analysis was conducted the same as NeuroNet-powered survival analysis, except the NeuroNet score was not used for prioritizing noncoding variants.

### Transcriptome analysis for the AnswerALS cohort

For AnswerALS dataset, RNA-seq data of iMNs and phenotype data were obtained for 245 ALS patients. Gene expression was normalized for gene length and then sequencing depth to produce transcripts per kilobase million (TPM) using Kallisto v0.46.0^78^. Read counts were trimmed Mean of M-values (TMM) normalized^79^ to account for differences in library size. For each gene, based on the expression level in and outside the top 25%, patients were split into two groups – high-expression and low-expression groups. We used multivariable linear regression to determine the relationship between gene expression level and rate of change in the ALSFRS-R, and Cox proportional-hazards regression on two patient groups to determine the relationship between gene expression and survival time from symptom onset. In both cases, age of disease onset, sex, site of disease onset, and *C9orf72* status were included in the model as covariates. Significance testing was performed for all genes expressed in iMNs as determined by >0 read counts in more than half of samples, and mean read counts across all samples >25th percentile. The ALSFRS-R was scored from a maximum of 48 where a reduced score indicates increasing disability^69^. We calculated the rate of change in the ALSFRS-R (i.e., Δ(ALSFRS-R)) assuming a linear decline from a score of 48 at symptom onset.

### CRISPR-editing of survival-associated genetic variant

The variant chr7:76,009,472:C>T (hg19 genome build, ENSEMBL) was introduced to *C9orf72*-ALS 183 iPSCs (passage 21) (RRID:CVCL_UF85) using CRISPR/Cas9-mediated homology directed repair (Synthego^80^). Donor sequence: GTGCCAGTCGCAGGCTCTCCAATCAGGAGGCTGCCGTGGGGCCAGGGGTGTGGCTCCGGTTCCCACGCGGGGCGAATTCCCTCTCCCGGTAGTCTCCCCAG. gRNA sequence:

GGGAACCGGAGCCGCGCCCC. *C9orf72*-ALS 183 iPSCs were derived from a 51 year old Caucasian male with *C9orf72* G4C2 repeat expansion and a diagnosis of familial ALS at the time of sampling^81, 82^. Editing efficiency was confirmed via Sanger sequencing using the following primers: fwd (5’-3’): CAGAAGCCCTGAGGAGTGC, rvs (5’-3’): GCCTCGGAGGTTTTTCTGGA. Sequencing traces were uploaded to the Synthego ICE Analysis tool (v3) (https://ice.synthego.com), and the indel efficiency within the mixed pool was calculated at 91%. Editing efficiency remained stable throughout motor neuron differentiation. Sequence traces are available on request. CRISPR-edited cells were engineered by Synthego (EditCo Bio).

### Tissue culture of iPSCs and iMNs

iPSCs were cultured in mTesR Plus media (StemCell Technologies) in Matrigel-coated (8.7 ug/cm²) 6-well plates (Corning) and maintained at 37°C, 5% CO2. Cells were passaged when ∼80% confluent using ReLeSR (StemCell Technologies), according to the manufacturer’s instructions. iPSCs were differentiated into neural progenitor cells (NPCs) using a modified version of the dual SMAD inhibition protocol^21, 83^. On the day after plating (day 1), after the cells had reached ∼100% confluence, the cells were washed once with PBS and the medium was replaced with neural induction medium (50% of KnockOut DMEM/F-12, 50% of Neurobasal), 0.5x N2 supplement (ThermoFisher), 1x Gibco GlutaMAX Supplement (ThermoFisher), 0.5x B-27 supplement (ThermoFisher), 50 U ml−1 penicillin and 50 mg ml−1 streptomycin, supplemented with SMAD inhibitors (DMH-1 2 μm; SB431542 10 μm; and CHIR99021 3 μm). This medium was changed every day for 6 days, and on day 7 the medium was replaced with neuralization medium supplemented with DMH-1 2 μm, SB431542 10 μm and CHIR 1 μm, All-Trans Retinoic Acid 0.1 μm (RA), and Purmorphamine 0.5 μm (PMN). The cells were kept in this medium until day 12 when it was possible to observe a uniform neuroepithelial sheet. At this point the cells were split 1:4 with Accutase (Gibco), onto matrigel substrate in the presence of 10 μm ROCK inhibitor (Y-27632 dihydrochloride, Tocris), giving rise to a sheet of neural progenitor cells (NPC). After 24 hours of incubation the medium was changed to neural medium supplemented with RA 0.5 μm and PMN 0.1 μm, and the medium was changed every day for 6 more days. On day 19 motor neuron progenitors were split with accutase into plates pre-coated with poly-l-ornithine (10 µg/ml for 1hr at RT followed by 3x washes in dH₂O) followed by matrigel (8.7 ug/cm² for 1hr at RT), and the medium was replaced with motor neuron differentiation medium supplemented with RA 0.5 μm, PMN 0.1 μm, compound E 0.1 μm (StemCell Technologies), BDNF 10 ng/mL, CNTF 10 ng/mL and IGF-1 10 ng/mL (Tocris) until day 28. On day 29, the media was replaced with neuronal maturation media (neurobasal media supplemented with 1% of B27, BDNF 10 ng/mL, CNTF 10 ng/mL and IGF-1 10 ng/mL). The cells were then fed every 72 hrs until day 40.

### Treatment of cells with antisense oligonucleotides (ASOs)

iMN monocultures were treated with 5 µM of either a scrambled control (GCGACTATACGCGCAATATG) or CCDC146-targeting (TGCTGACTAATTGTGGTTAC) ASO (Integrated DNA Technologies) in neuronal maturation media for 72 hours prior to downstream assessment of ciliary function by immunocytochemistry or immunoblotting.

### Measurement of gene expression in iMNs

Gene expression was determined via quantitative real-time polymerase chain reaction (qRT-PCR). Primer sequences were as follows: *FGL2: Fwd: TCAAAGTGTCCCAGCCAAGAA, Rvs: ACTCTGTAGGTCTCACTGCTTC. FAM185BP: Fwd: TCAGGGTCTGGCTGTGTAGA, Rvs: TCCCAGACAGATCACTTTGCC. CCDC146: Fwd: ACTCCACAAAGCTCATCAGAAAG, Rvs: TCCTGAAGTTTTGATACAATTTTGC.* RNA was extracted using a Direct-zol RNA Purification Kits (Zymo) according to the manufacturer’s instructions. 2μg of total RNA was converted to cDNA in an initial reaction mixture containing 1μL 10mM dNTPs, 1μL 40μm random hexamer primer (ThermoFisher Scientific), and DNAse/RNAse-free water to a total volume of 14μL. This mixture was heated for 5 minutes at 70°C then incubated on ice for 5 minutes. 4μL of 5x FS buffer, 2μL 0.1M DTT, and 1μL M-MLV reverse transcriptase (200U/μL) (ThermoFisher Scientific) were then added, and cDNA conversion was performed in a PCR thermocycler (37°C for 60 minutes, 85°C for 10 minutes). cDNA was amplified using RT-PCR with Brilliant III SYBR Green (Agilent) as per the manufacturer’s instructions. Ct analysis was performed using CFX Maestro software (BioRad). β-actin was chosen as a reference gene because expression is stable in iPSC-derived neurons^84^. qPCR products were sequenced to confirm target specificity and sequence traces are available on request. Statistical analysis was performed using GraphPad Prism version 10.0.0.

### Longitudinal survival tracking of iMNs

Survival of iMN was assessed from day 28-40 using an Incucyte® Live Cell Analysis System (Sartorius) at 20x magnification across 3 biological repeats. Neurite length (mm/mm^2^) was used as a readout for cell survival. For each biological repeat, 10 wells were imaged per group with 9 fields of view per well. The neurotrack module (Sartorius) was used to determine neurite metrics from brightfield images. In each case statistical significance was calculated by applying a paired *t*-test at each timepoint. Statistical analysis was performed using GraphPad Prism version 10.0.0.

### Immunocytochemistry

For immunostaining, neural progenitor cells (NPC) and motor neurons (MN) were washed with phosphate-buffered saline (PBS) and fixed with 4% paraformaldehyde for 10 min at room temperature. After fixation samples were washed three times with PBS and permeabilized with 0.3% Triton X-100 diluted in PBS for 5 min. The cells were subsequently blocked in 5% Donkey serum (Millipore) for 1h at room temperature (RT). After blocking, cells were incubated with the appropriate primary antibodies: mouse anti-nestin 1:200 [Abcam #ab22035]; rabbit anti-Pax6 1:50 [Abcam #ab5790]; guinea pig anti-MAP2 1:1000 [Synaptic Systems #188004]; chicken anti-TUJ1 1:1000 [Merck Millipore #AB9354]; mouse anti-SMI32 1:1000 [Biolegend #801701]; rabbit anti-HB9 1:250 [Abcam #ab221884]; goat anti-ChAT 1:50 [Merck #AB144P]; rabbit anti-CCDC146 [Atlas Antibodies #HPA020082] diluted in PBS containing 5% of DS overnight. Cells were then washed with PBS three times. Fluorescent secondary antibodies (Alexa Fluor 488, 561, or 647, diluted 1:1000 with 5% DS) (ThermoFisher Scientific #A-21202, #A-21432, A-21450, #A-32744, #A-21206) were subsequently added to the cells and incubated for 1h at RT. The samples were washed with PBS three more times and incubated with Hoechst 33342 (Invitrogen) for nuclear staining for 5 minutes. For primary cilium staining, a CoraLite® Plus 488-conjugated ARL13B polyclonal antibody (Proteintech #CL488-17711) was applied for 1hr after all washes to limit cross-reactivity with other primary antibodies of the same species. All experiments included cultures where the primary antibodies were not added; non-specific staining was not observed in these negative controls. Images were obtained using an Opera Phenix™ High Content Screening System (Revvity) at 40x magnification to generate a high-resolution z-stack at 0.5 µm intervals throughout the entire nuclear volume of the cell. Images were analyzed using the Harmony™ Image analysis system. 488, 561, and 647 nm lasers along with the appropriate excitation and emission filters were used. These settings were kept consistent while taking images from all separate differentiations.

### Measurement of cilia length

CIlia length was the longest dimension of cilia immunostaining measured in maximum projection through visualised cilia. Each motor neuron has only one cilium. A technical replicate was taken to be the mean cilium length in a single field of view from at lest three separate motor neuron differentiations per condition. A paired t-test was used to determine statistical significance and account for variability between motor neuron differentiations.

### Measurement of microtubule dynamics

iPS-MN were transfected with 1 µg EB3_mScarlet (Addgene #98826, kindly gifted from the Dorus Gadella lab) at day 39 using Lipofectamine™ 3000 (ThermoFisher Scientific), according to the manufacturer’s guidelines. For live imaging by TIRF microscopy 24 hours post transfection, cells were cultured in phenol red-free neurobasal media supplemented with 10mM Hepes, 1% B-27 supplement, and imaged using a custom-built Nikon microscope with an iLas2 multi-application unit (Gataca Systems). Time-lapse imaging of EB3 comets within axons was performed at 200 ms intervals for 2 minutes using a 568 nm laser. Axons were straightened with the segmented line tool in FIJI, resliced, and then converted into kymographs using a maximum intensity z-projection. The velocity of EB3 comets was analysed using the FIJI plugin, TrackMate^85^.

### Western blotting

iMN were lysed on ice in IP-lysis buffer (50 mM Hepes pH7.5, 150 mM NaCl, 10% glycerol, 0.5% Triton X-100, 1 mM EDTA, 1 mM DTT, protease inhibitor cocktail (Sigma)), passed 10x through 25-guage needles and incubated on ice for 10 minutes. Samples were centrifuged at 17,000 xg for 5 minutes at 4°C. Protein extracts were quantified using Bradford Reagent (BioRAD), resolved by SDS–PAGE (15% resolving gel; 5% stacking gel), and electroblotted onto nitrocellulose membranes. Membranes were blocked in 5% BSA + TBST (20 mM Tris, 137 mM NaCl, 0.2% (v/v) Tween® 20, pH 7.6) for 1 hour at room temperature (RT). Membranes were probed using the relevant primary antibody [anti-CRABP1 (1:1000 dilution), Proteintech #12588-1-AP; anti-TUJ1 (1:1000 dilution), Abcam #ab18207] diluted in 5% BSA + TBST overnight at 4°C. Membranes were washed 3x in TBST, incubated with an Alexa Fluor 680-AffiniPure Donkey Anti-Rabbit IgG (H+L) secondary antibody (Jackson #711-625-152) diluted in TBST for 1 hour at RT, and washed 3x in TBST. Fluorescence signal intensity was analysed using the Odyssey® Fc Imaging System (LI-COR Biosciences).

### Generation of iPSC-derived 3D-induced neurons

To generate iPSC-derived 3D-induced neurons (3D-iNs), iPSCs were first co-transfected (Lipofectamine Stem, Thermofisher) with a piggybac expression cassette containing a dox-inducible h*NGN2* transgene and mApple fluorescent reporter (addgene 182308) and the Super piggyBac Transposase expression vector (SBI). After a brief (24h) recovery, transfected iPSCs were replated using accutase+10μm ROCK-inhibitor (Ri, Selleckchem) at sparse densities where an initial selection with 20μm blasticidin (Cayman, BSD) was performed. BSD concentrations were increased until a highly pure population of mApple fluorescent cells were observed and subsequently used for downstream studies. To generate the 3D-iN cultures, iPSCs expressing the h*NGN2* piggybac construct were dissociated with accutase and 10μm ROCK-inhibitor (Selleck) and seeded into Matrigel coated 6-well plates at ∼200,000 cells per well in induction medium containing DMEM+Glutamax (Thermofisher), NEAA (Gibco), 1% penicillin/streptomycin, N2 supplement (Gibco), and 10ng/ml each of BDNF (R&D) and NT-3 (Peprotech) plus doxycycline (2μg/mL) to drive the expression of the h*NGN2* cassette. 48 hours after induction, induction medium was replenished and BSD (20-40μg/mL) was added if non-converting, non-transgenic cells were observed. 72 (Day 3) hours after induction, BrdU (Millipore Sigma) at 40μm was added to eliminate dividing cells. By Day 5 of induction, the pure population of converting iNs were replated using accutase and Ri (20μM) to ultra-low attachment 96-well U-bottom plates (Corning) containing neuronal maintenance medium (MM): neurobasal (Thermofisher), N2 and B27 supplements (Gibco), 1%penicillin/streptomycin, glutamax (Thermofisher, 100x), and NEAA (Thermofisher, 100x) plus BDNF and NT-3 (10ng/mL each) at densities of ∼50,000-80,000 neurons per well, where the cells would coalesce into 3D-iN cultures. 3D-iNs were maintained in this format until Day 40 for survival tracking with medium changes (MM plus BDNF and NT3) every 3 days.

### 3D-iN longitudinal survival tracking

3D-iNs were allowed to mature until Day 30 after dox and were then transduced with a *SYN1*::GFP lentivirus to enable labeling of individual neurons as previously described^45, 48^. 72-hours prior to the start of the survival assay (Day 37 after Dox), 3D-iNs were treated with ASO gapmers designed using an in-house strategy; 10μm of either a scrambled or *CCDC146* ASO were administered via gymnotic uptake. On Day 39 (24 hours prior to start of tracking), 3D-iNs were embedded in 100% Matrigel (Corning) for 30 minutes to allow gelling. MM was replenished after the 3D-iNs were fixed. Following a baseline image at Day 0 of the tracking (Day 40 of differentiation), glutamate (L-Glutamic acid monosodium salt hydrate; 5mM Milipore) was added to the medium and replenished every other day thereafter. Continuous z-stacks starting from the 3D-iN surface were captured every other day (from Day 0) using a Zeiss AxioZoom.v16 wide-field upright fluorescence microscope. Image processing and analysis were performed using ImageJ software (NIH). To quantify neuronal death ∼100 neurons were tracked across 4 replicates per line per condition. Hazard ratios (HR) were calculated using the log-rank method; for each replicate across each condition, the HR was calculated as a comparison to the aggregate survival of the scrambled ASO control spheroids.

### hTDP43 mouse studies

Mouse strains C57BL/6J and B6;SJL-Tg(Thy1-TARDBP)4Singh/J (stock number: 012836) were purchased from the Jackson Laboratory. The murine Thy1 promoter drives expression of human TDP-43 in neurons and other CNS/PNS cells. Animals were housed in a temperature and humidity-controlled environment under standard conditions with access to food and water ad libitum. Animals were subjected to 12-hour light/dark cycles. All animal care and experimental procedures were performed in accordance with local institution guidelines of the University of Southern California and approved by the Institutional Animal Care and use Committee of the University of Southern California (protocol number: 11938).

hTDP43 transgenic mice were purchased from the Jackson Laboratory (stock number: 012836). hTDP43^Tg/Tg^ homozygous mutant mice were generated by crossing hTDP43^Tg/+^ heterozygous mice with hTDP43^Tg/+^ heterozygous mice. Genotyping primers are as follows: Common primer: (5’- TGAAATCCGGGTGGTATTGG-3’); WT primer: (5’- GGTGAGTTTAACCTTCAAGGGCT-3’); Transgene primer: (5’- AGCTTGCTAGCGGATCCAGAC-3’). Homozygous transgenic mice were identified by a single, 500 bp band, heterozygotes with a 303 bp and 500 bp bands, and wild-type (WT) mice with a single 303 bp band. Homozygous transgenic animals and their WT littermate controls were used for the subsequent ASO studies.

### ASO mouse treatments

25µg of a negative control or Ccdc146 ASO were administered by intracerebroventricular (ICV) injection at post-natal day 1 (P1). ASOs were ordered from Integrated DNA Technologies and were designed with chemical modifications to improve their potency and efficiency for gene knockdown. Phosphorothioate linkages were used within the sequence to provide resistance to nucleases, the 2’-O-methoxy-ehtyl (MOE) modification was added to enhance nuclease stability and binding affinity for the target RNA. ASOs were ordered with additional HPLC and sodium/salt exchange purification steps to remove potentially toxic byproducts during synthesis. Purity of the ASOs was confirmed by IDT via mass spectrometry. ASO sequences used in these studies are as follows (m=2’-O-Methyl, *-phosphorothioate linkage, MOE = 2’-Methoxyethyl): msNC ASO: /52MOErC/*/i2MOErC/ /i2MOErT/ /i2MOErA//i2MOErT/ A*G*G*A*C*T*A*T*C*C*/i2MOErA/ /i2MOErG/ /i2MOErG/* /i2MOErA/*/32MOErA/; msCcdc146 ASO: /52MOErC/*/i2MOErA/ /i2MOErA/ /i2MOErT/ /i2MOErC/ T*G*C*T*G*C*T*C*C*A*/i2 MOErC//i2MOErT/ /i2MOErA/* /i2MOErA/*/32MOErC/

### ICV injection procedure

D1 neonatal mice were anesthetized by placing them on a paper towel lined bucket of ice to induce hypothermia. The injection site targeting the lateral ventricles was defined as 2/5 the distance from the anatomical landmark, lambda to the eye. To perform the ICV injection, a 900 series Hamilton syringe (10µL) adapted with the bottom portion of Neuros model 1701 RN syringe. A 33-gauge needle was inserted 2mM deep, perpendicular to the skull surface into the lateral ventricle (left). Mice received 3µL of the respective NC ASO or Ccdc146 ASO in PBS (25µg). Following the injection, mice were placed under a heat lamp in a new cage containing soiled bedding from the parental cage and were monitored until they had fully recovered. Motor phenotyping (gait, kyphosis, and tremor) was examined starting at P14 and recorded every other day until D22. Phenotype score was determined according to a rubric previously reported (PMID: 28405022). A higher score denotes a more severe phenotype (0=no phenotype, 4=most severe/end stage). The humane euthanasia endpoint was established to be when the mouse was no longer able to right itself. Hydrogel, powdered pellets, and high caloric diet gel were placed on the bottom of each cage containing the mutant mice.

### Mouse tissue collection

WT or hTDP43^Tg/Tg^ were treated with a NC ASO or Ccdc146 ASO at postnatal day 1 via ICV injection. For tissue collection for, D22 mice were anesthetized and intracardiac perfusion was performed first with PBS and then with 4% PFA in PBS, the tissue was then fixed in 4% PFA for another 4 hours. Tissue was then incubated in 15% sucrose in PBS+.01% sodium azide for 24 hours and then 30% sucrose in PBS +.01% sodium azide for another 48 hours prior to embedding and immunofluorescence analysis. For RNA isolation, mice were euthanized and isolated brains were separated into two hemispheres, one hemisphere and spinal cords were lysed for RNA analysis and the other was frozen at -80°C.

### Mouse histology

Cryoprotected tissues were embedded in O.C.T (Fisher, cat: 23730571) and cryosectioned into 16µM slices using a Leica CM3050S cryostat onto Superfrost Plus Microscope slides (Fisher) and stored at -80°C until use. For spinal cord neuron counts, lumbar spinal cord sections were rehydrated in 10x PBS for >40 minutes and washed in PBS+.1% Triton X-100 for 10 minutes. Slides were then washed in PBS for 2, 5-minute washes. The slides were then incubated with NeuroTrace (Fluorescent Nissl Stain, Thermo N2148) diluted 1:50 in PBS for 20 minutes in the dark. After removing the stain, slides were washed for another 10 minuites in PBS+.1% Triton-X-100 for 10 minutes, 2x5 minute washes in PBS and 2 hours at room temperature/overnight at 4°C in PBS prior to mounting. To quantify the neurons in the ventral horn region of the spinal cord, 3 sections per animal were stained with NeuroTrace and image using a Zeiss LSM 800 microscope. The lateral motor column was used for analysis. The counts were quantified based on the average total neuron counts in the lumbar hemicords across all 3 sections per animal, per treatment condition. To quantify the cellular localization of TDP43/tdp43, the nuclear and cytoplasmic fluorescence intensity was measured using an antibody with reactivity for both murine and human protein (10782-2-AP, Proteintech, 1:400). The fluorescence intensity of the TDP43 staining in the cell nucleus and cytoplasm were measured using Image J and recorded as the nuclear to cytoplasmic ratio. TUJ1+ motor neurons in the ventral spinal cord were imaged using Zeiss LSM 800 with oil immersion at 63x. 15–25 neurons were quantified per animal.

### Small-molecule drug search

We searched chemical activity databases ChEMBL^86^ and PubChem^87^ for CCDC146 and its similar proteins, but did not obtain any active compounds for these proteins. Here, similar proteins were defined as those with at least 50% sequence identities to the canonical sequence of human CCDC146 protein, provided by the UniProt database^88^. We also searched patent databases including WIPO (https://patentscope.wipo.int/), EPO (https://www.epo.org/en/searching-for-patents) and USPTO (https://www.uspto.gov/patents/search), using search terms “CCDC146” and “coiled-coil domain-containing protein 146”, to confirm that there is currently no available drugs for this target.

### Protein structure and binding site prediction

The predicted protein structure of CCDC146 protein was obtained from the AlphaFold Protein Structure Database^89^ (https://alphafold.ebi.ac.uk/). To evaluate the druggability of CCDC146, we predicted potential ligand binding sites on its surface and obtained negative prediction results using P2Rank^67^, which is an accurate ligand binding site prediction tool.

## Supplementary Figures

**Supplementary Fig. 1.**
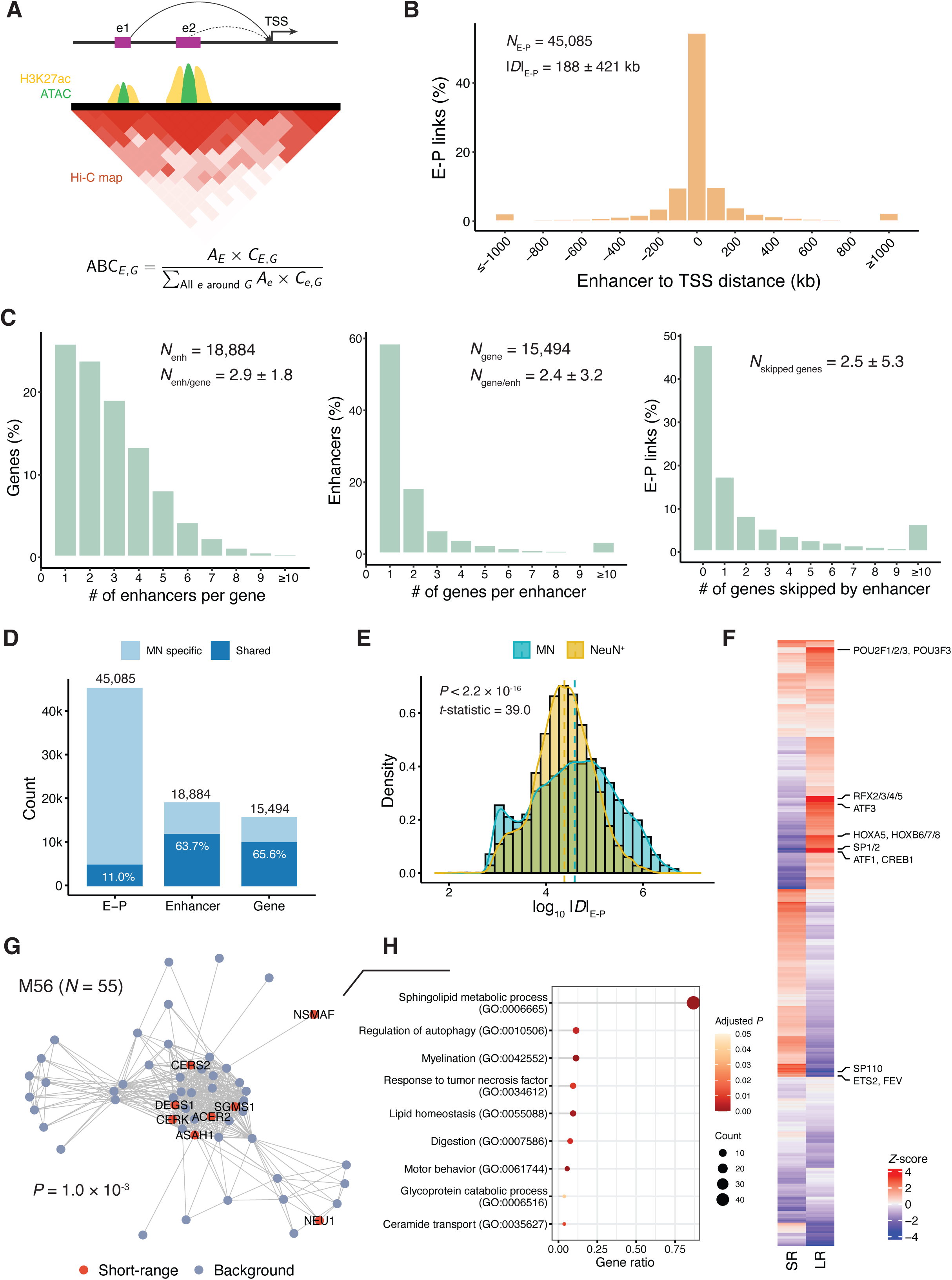
Enhancer-promoter (E-P) interactions in human motor neurons. (A) Graphical illustration of the ABC model. TSS, transcription start site; E, enhancer; G, gene. (B) Distribution of distance from enhancer to TSS. (**C**) Statistics of E-P links, including number of enhancers per gene, number of genes per enhancer, and number of genes skipped by enhancer. enh, enhancer. (**D**) Overlap between E-P links in MNs and those in neuronal cells (NeuN^+^). (**E**) Distribution comparison of E-P links between MNs and neuronal cells. Density curves were added. Dashed vertical lines represent distribution medians. *P*-value by two-side *t*-test. (**F**) Differential motif enrichment between long-range and short-range E-P links. LR, long-range; SR, short-range. (**G**) Network module enriched with genes targeted by short-range E-P links. *P*-value by hypergeometric test. (**H**) Gene ontology (GO) enrichment for M56 genes. Redundant GO terms were removed using the “simplify” function provided by “clusterProfiler”. GO terms with adjusted *P* < 0.1 were visualized. *P*-value by two-sided Fisher’s exact test.

**Supplementary Fig. 2.**
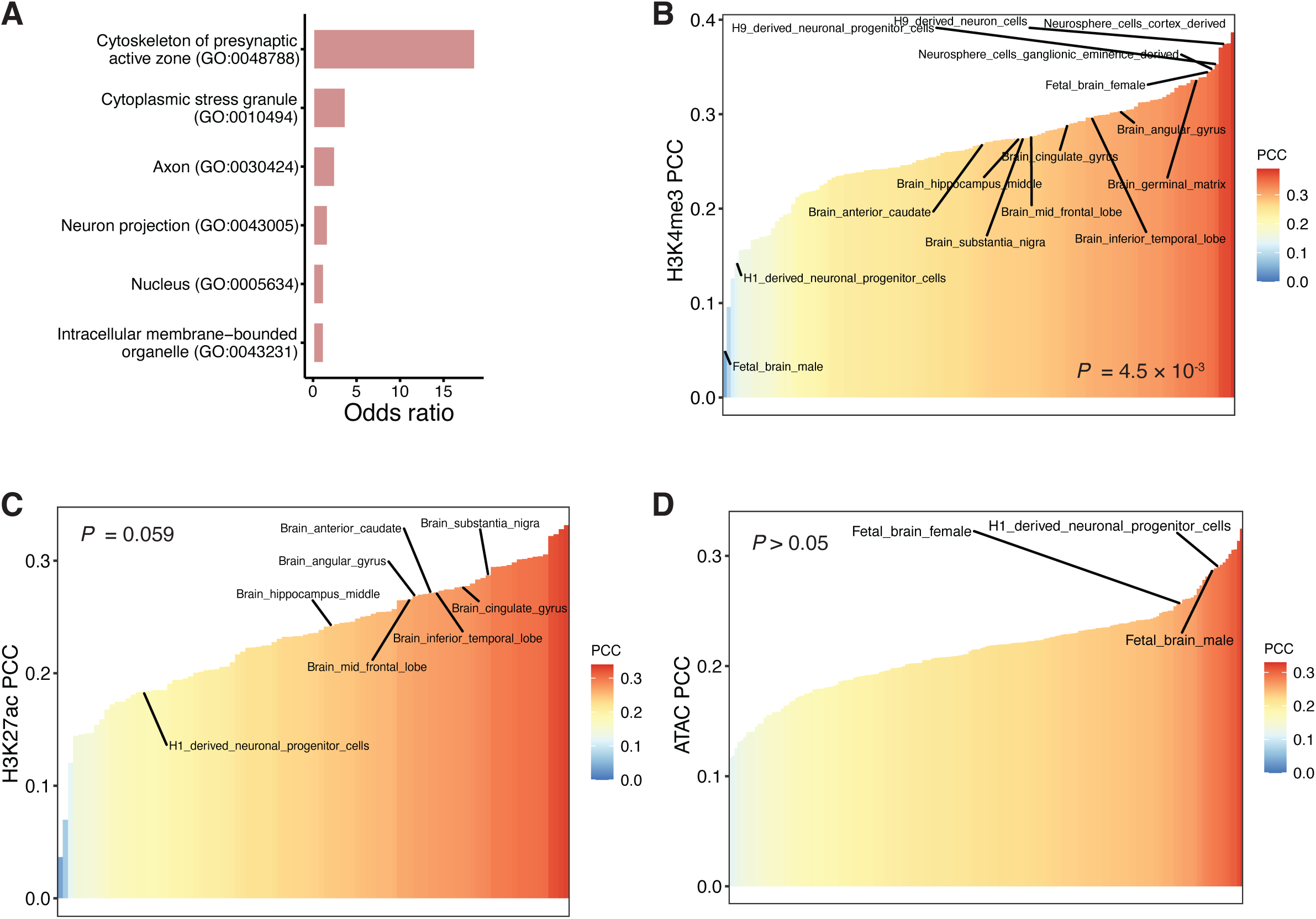
Additional results for E-P links and NeuroNet prediction. (**A**) Gene ontology (GO) enrichment for genes with short-range enhancers in iPSNs. GO terms with adjusted *P* < 0.1 (BH correction) were shown. *P*-value by two-sided Fisher’s exact test. (**B-D**) Pearson correlations between NeuroNet and DeepSEA predictions for H3K4me3 (**B**), H3K27ac (**C**), and ATAC (**D**). *P*-value by two-sided Fisher’s exact test. Neuron- and brain-related tissues and cell types were highlighted. PCC, Pearson correlation coefficient.

**Supplementary Fig. 3.**
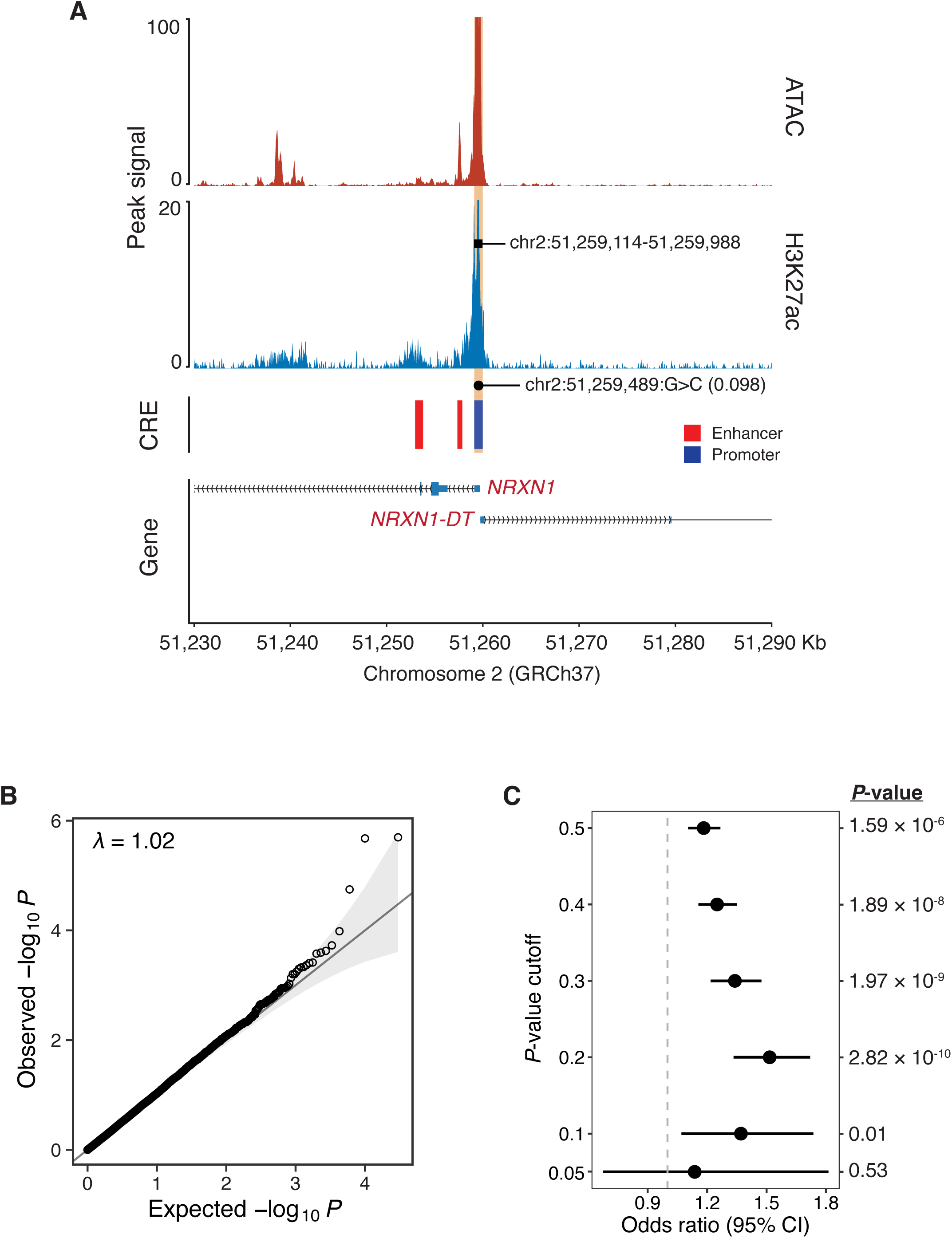
Additional results for rare variant burden analyses. (**A**) Regulatory relationships between chr2:51,259,489:G>C and target genes. Peak signals from MNs derived from CS14-iPSCs were visualized. Significant genes in NeuroNet-informed survival analysis were highlighted in red. CRE, cis-regulatory element. (**B**) Quantile-quantile (QQ) plot of a standard rare variant burden-based survival analysis without NeuroNet-based variant filtering. Genes sharing the same set of variants were collapsed into groups. (**C**) Overlap of genes between survival and risk analysis with different *P*-value cutoffs. Overlap was examined by two-sided Fisher’s exact test. The dot and error bar indicate the odds ratio and 95% CI, respectively. The dashed gray line represents odds ratio = 1. CI, confidence interval.

**Supplementary Fig. 4.**
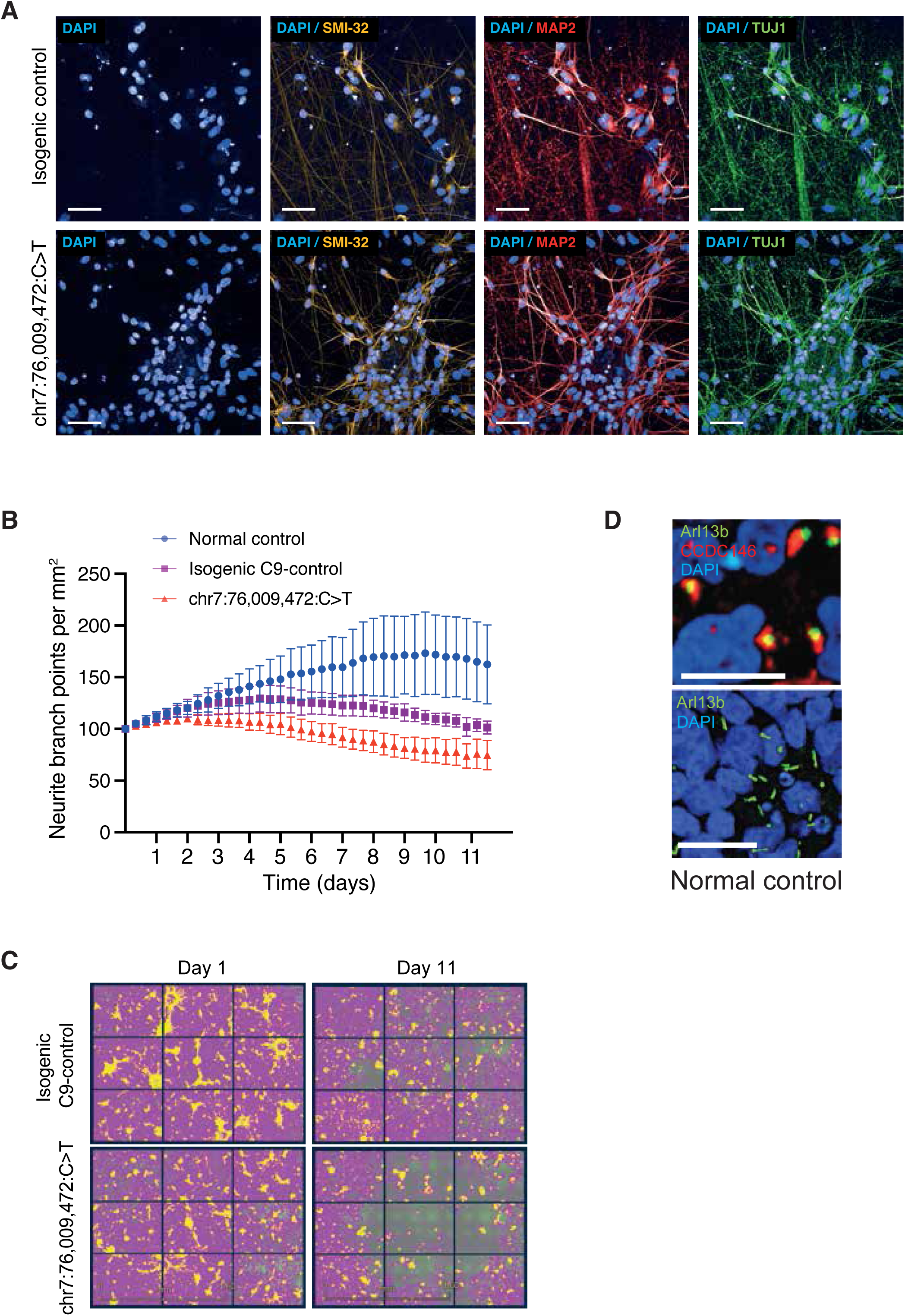
Additional results for CRISPR/Cas9-mediated modeling of chr7:76,009,472:C>T in iPSC-derived *C9orf72*-ALS MNs. (**A**) Representative images of iPSC-derived MNs (iMN). Top panels: isogenic control; bottom panels: edited. iMNs were studied at day 40 post-iPSC-stage when they expressed a battery of neuronal markers, including SMI-32, MAP2, and TUJ1. Scale bar: 50μm. (**B**) Quantification of the number of neurite branch-points in an age-and sex-matched normal control, an isogenic control carrying a G4C2-repeat expansion of *C9orf72*, and chr7:76,009,472:C>T-edited iMNs across 12 days of tracking using an Incucyte live-cell analysis system (Sartorius). The dot and error bar represent the mean and standard error, respectively. (**C**) Representative images from day 1 and day 11 for isogenic control neurons carrying a G4C2-repeat expansion of *C9orf72* (top panels) and chr7:76,009,472:C>T-edited neurons (bottom panels). Cell soma are coloured yellow and neurites are coloured purple. Scale bar 1.6mm. (**D**) Immunocytochemistry for CCDC146 and primary cilia stained with Arl13b in iMNs derived from a neurologically healthy control. Scale bar: 50μm.

**Supplementary Fig. 5.**
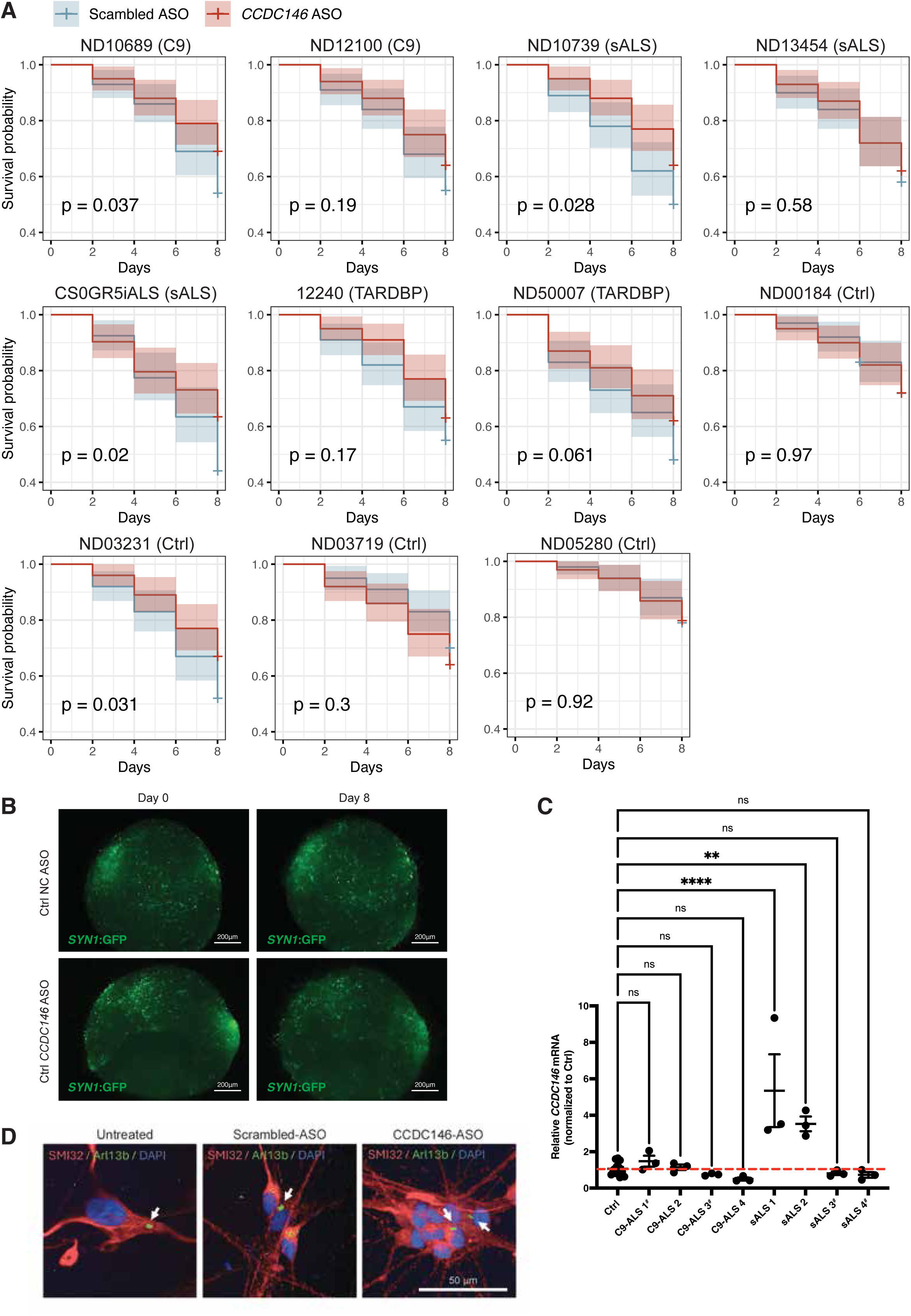
Additional results for ASO suppression of *CCDC146* in ALS patient-derived neurons. (**A**) Kaplan-Meier (KM) estimator plots of 3D-iN survival starting (day 0) from day 40 after initiation of iPSC differentiation. The shaded area represents 95% confidence interval (CI). *P*-value by log-rank test. C9, *C9orf72*; sALS, sporadic ALS; ctrl, control. (**B**) Representative images of control iNs treated with the scrambled ASO (top panels) or the *CCDC146* ASO (bottom panels). Images were taken from approximately the same location on the plat at Day 0 (left panels) and Day 8 (right panels) showing no obvious toxicity. Scale bar: 200μm. Ctrl, control. (**C**) Quantification of *CCDC146* mRNA expression via q-RT-PCR in *n* = 1 control line and *n* = 8 ALS patient-derived iN lines. *n* = 3 technical replicates per cell line. The central bar and error bar represent the mean and standard error, respectively. The red dashed line indicates control expression level. *P-*value by the Dunnett’s test. **, adjusted *P* < 0.01; ****, adjusted *P* < 0.0001; ns, not significant; ^#^, patient cell lines used in ASO experiments. (**D**) Immunocytochemistry for primary cilia stained with Arl13b across different treatments. Scale bar: 50μm.

**Supplementary Fig. 6.**
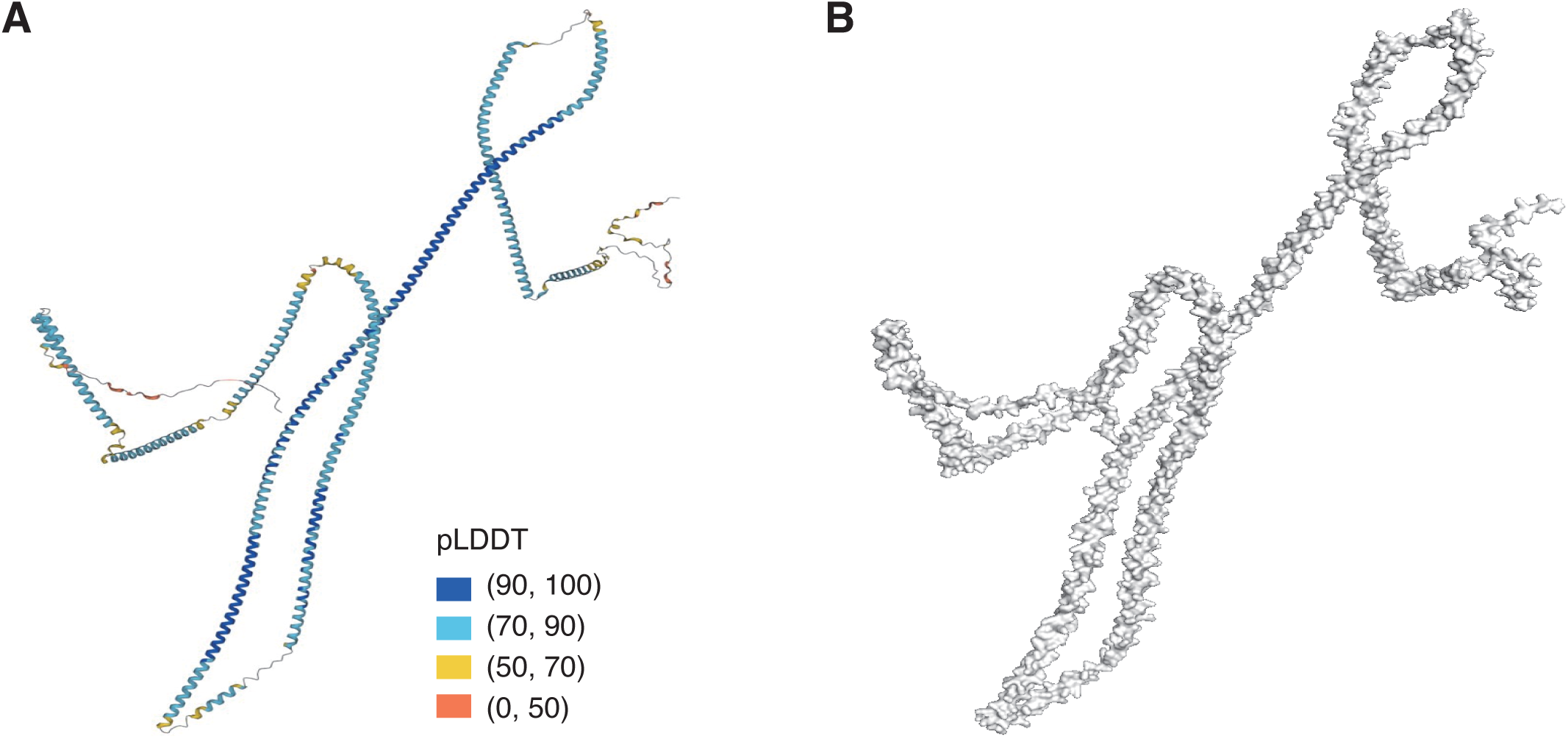
The AlphaFold-predicted three-dimensional structure of CCDC146 protein. (**A**) The carton illustration of the predicted protein structure of CCDC146, obtained from https://alphafold.ebi.ac.uk/entry/Q8IYE0. pLDDT, predicted local distance difference test. (**B**) Protein surface of the predicted structure, illustrated using PyMol (https://pymol.org/).

**Supplementary Fig. 7.**
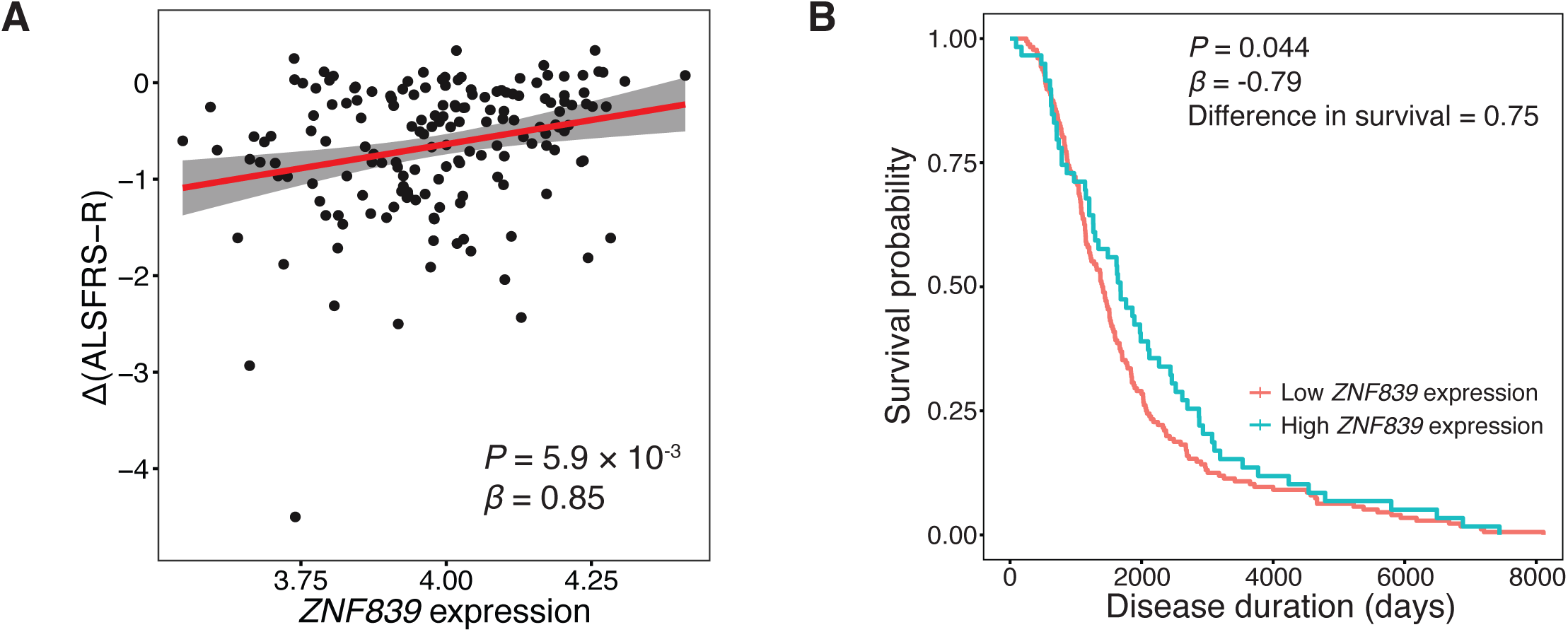
Relationship between *ZNF839* expression and clinical markers of disease progression. (**A**) Correlation between the expression of *ZNF839* and rate of disease progression as measured by clinical symptoms. The ALSFRS-R is scored from a maximum of 48 with a reduction in the score signifying increased disability. The red line indicates the regression line; the gray area represents the standard error. *P*-value by multivariable linear regression with sex, age of onset, site of onset, and *C9orf72* status as covariates. (**B**) Kaplan-Meier (KM) estimator plots of patients with high and low *ZNF839* expression. Gene expression level in the top 25% was defined as “high expression” (*n* = 59), otherwise as “low expression” (*n* = 176). The difference in mean survival between two groups was 0.75 years. *P*-value by Cox proportional-hazards (PH) regression.

## Supplementary Tables

**Supplementary Table 1.** Enhancer-promoter links in iPSC-derived motor neurons

**Supplementary Table 2.** Summary statistics of SKAT-O test for ALS risk

**Supplementary Table 3.** Summary statistics of NeuroNet-powered rare variant burden test for ALS survival

**Supplementary Table 4.** Summary statistics of rare variant burden test for ALS survival without variant filtering

**aSupplementary Table 5.** Clinical phenotypes of sequenced ALS patients carrying survival-associated rare noncoding variants

**Supplementary Table 6.** Clinical phenotypes of patient iPSC lines

