## Supplementary Results for "Antisense oligonucleotide depletion of *CCDC146* is a broad-spectrum therapeutic strategy for ALS"

***Constructing the regulome in human motor neurons***

To systematically study transcriptional regulation in human motor neurons, we mapped MN enhancer-promoter (E-P) interactions by applying the activity-by-contact (ABC) model^1^ (**Supplementary Fig. 1A**; **Methods**) to the transcriptomic (measured by RNA-seq) and epigenomic (measured by ATAC-seq, H3K27ac ChIP-seq, and Hi-C) profiling data for iMNs derived from neurologically normal donors^2^ (*n* = 3). We identified 45,085 E-P interactions for 15,494 expressed genes (absolute E-P distance: mean = 188 kb, standard deviation (std) = 421 kb; **Supplementary Fig. 1B** and **Supplementary Table 1**). We identified at least one non-promoter cis-regulatory element (CRE) for 85.1% of expressed genes. Of all expressed genes in iMNs, 74% were associated with more than one enhancer (mean = 2.9, std = 1.8; **Supplementary Fig. 1C**). Similarly, 41% of enhancers were predicted to regulate more than one target gene (mean = 2.4, std = 3.2; **Supplementary Fig. 1C**). This multi-to-multi relationship between enhancers and target genes highlights the complexity of transcriptional regulation within MNs. Importantly, 52% of E-P links skipped the closest gene (mean number of skipped genes = 2.5, std = 5.3; **Supplementary Fig. 1C**), indicating the importance of long-range gene regulation within MNs.

To assess the specificity of MN E-P links, we compared E-P links between MNs and other neuronal cells based on a recent study^3^. While the majority of enhancers (63.7%) and expressed genes (65.6%) in MNs overlapped with those in other neuronal cells, a large proportion of MN E-P links (89.0%) were cell-type-specific (**Supplementary Fig. 1D**). This result highlights the difference of transcriptional regulation between MNs and other neuronal cells, emphasizing the necessity for focused study of MNs. Interestingly, the distance of E-P links in MNs tended to be longer than those in other neuronal cells (median: 39 kb (MN) vs. 24 kb (NeuN^+^); *P* < 2.2 x 10^-16^, two-sided *t*-test; **Supplementary Fig. 1E**).

To provide a functional characterization of gene regulation in MNs, we performed transcription factor (TF) binding motif analysis within enhancers. We compared TF binding motifs between short-range (<5% absolute E-P distance, *n* = 945) and long-range (>95% absolute E-P distance, *n* = 945) enhancers (**Supplementary Fig. 1F**; **Methods**). The binding motif of SP110 was enriched in short-range enhancers; this TF has been implicated in the regulation of penetrance in an animal model of ALS^4^. TFs such as ATF1 and ATF3, whose binding motifs were enriched in long-range enhancers, are primarily involved in homeostasis in response to extracellular stimuli^5^. This is particularly important for cells such as MNs that are not only post-mitotic but also show limited functional redundancy because of their high energy demands and extreme axonal length. Similarly, the SP1 binding motif was enriched in long-range enhancers, and SP1 has been implicated in maintaining neuronal survival in the context of oxidative stress^6^.

Next, we examined the biological implication of E-P links from a gene perspective. As expected, the set of genes linked to enhancers via short-range interactions (<5% absolute E-P distance, *n* = 775) were enriched with biological functions common to all neuronal subtypes (adjusted *P* < 0.1, Benjamini-Hochberg (BH) correction, two-sided Fisher’s exact test; **Methods**) including ‘cytoskeleton of presynaptic active zone’ (GO:0048788) and ‘axon’ (GO:0030424). Genes with long-range enhancers (>95% absolute E-P distance, *n* = 774) were enriched with ‘metalloendopeptidase inhibitor activity’ (GO:0008191; adjusted *P* = 0.045, BH correction, two-sided Fisher’s exact test). Metalloendopeptidase acts extracellularly and it is notable that reduction in extracellular matrix (ECM) proteins exacerbates ALS neurodegeneration^7^. Moreover, the loss of metalloendopeptidase in neurons has been linked to impaired motor activities and cognitive deficits by altering axonal maturation and myelination in the central nervous system (CNS)^8^.

To provide a systems view, we further performed a network analysis (**Methods**) based on human protein-protein interactions^9^ (PPIs). We discovered one module (M56, *n* = 55; **Supplementary Fig. 1G**) which was significantly enriched with genes targeted by enhancers in short range (*n* = 8; *P* = 1.0 x 10^-3^, hypergeometric test). Interestingly, genes within this module were enriched with biological pathways that are fundamental to both MN function and ALS pathogenesis (adjusted *P* < 0.1, BH correction, two-sided Fisher’s exact test; **Supplementary Fig. 1H**), including neuromuscular function (GO0061744), autophagy^10^ (GO:0010506), and sphingolipid (GO:0006665) and ceramide transport (GO:0035627) which are important for organization of MN function and signaling^11^. No module was found to be enriched with long-range targeted genes. Altogether, our results reveal the complexity and specificity of gene regulation in MNs, highlighting the unique regulatory patterns for cis-regulatory elements (CREs) and genes of functional significance in MNs and ALS.
